# Suicidal thoughts and behaviours in Cape Town: a cross-sectional study of prevalence, social, contextual, and clinical correlates

**DOI:** 10.64898/2025.12.24.25342957

**Authors:** Mpho Tlali, Reshma Kassanjee, Leigh L van den Heuvel, Stephan Rabie, John Joska, Catherine Orrell, Soraya Seedat, Hans Prozesky, Kristina Adorjan, Mary-Ann Davies, Leigh F Johnson, Andreas D Haas

## Abstract

**Background:** Suicidal thoughts and behaviours (STBs) are traditionally attributed to mental disorders; however, increasing evidence indicates that social determinants contribute to suicide risk. We examined STBs and their socioeconomic, psychosocial and clinical correlates in two communities in Cape Town.

**Methods:** We conducted psychiatric diagnostic interviews (MINI) at three public-sector facilities (2023-2024). Adults aged ≥18 years, with and without HIV, were recruited in a 2:1 ratio. We examined associations between suicidal ideation and sociodemographic factors, violence exposure, perceived stress, mental disorders, and HIV, reporting average marginal effects (AME) as percentage-point differences in predicted probability.

**Results:** We enrolled 613 participants (63.9% female; 65.4% living with HIV; median age 39). The prevalence of past 30-day suicidal ideation was 14.0%, while 22.2% reported a lifetime suicide attempt. Suicidal ideation was more likely among females (8.52 percentage points, 95% CI 3.08–13.96), those reporting violence in the community (7.52, 0.87–14.17) or in the family (14.09, 4.56 –23.62), those with moderate (7.76, 2.76 –12.77) or high perceived stress (33.58, 20.67– 46.50), and those with depression (27.13, 14.67– 39.58), post-traumatic stress disorder (27.98, 12.12 – 43.84), or alcohol use disorder (8.34, 1.53 –15.15). Associations with high perceived stress and community violence persisted after adjustment for mental disorders. HIV status and other sociodemographic factors showed no evidence of association.

**Conclusion:** STB prevalence was high in peri-urban communities in Cape Town and strongly associated with mental disorders, violence exposure, and perceived stress. These findings underscore the role of structural and psychosocial stressors in shaping suicide risk in low-income communities.

## Introduction

Suicide is a major public health challenge (GBD 2021 Suicide Collaborators, 2025). The World Health Organization (WHO) estimates that approximately 700,000 people die by suicide each year and suicide is the third leading cause of death in young adults aged between 15 and 29 years (World Health Organization, 2021). Low- and middle-income countries (LMIC) bear the largest burden of suicide, accounting for nearly 80% of all suicide deaths (World Health Organization, 2021). South Africa is among the countries with the highest suicide rates worldwide, with an estimated rate of 22.3 per 100,000, corresponding to about 14,000 deaths annually (World Health Organization, 2021, 2025). The rate is more than three times higher in males (35.4 per 100,000) than in females (9.9 per 100,000) (World Health Organization, 2025).

Suicide can be conceptualized as progression from thinking about ending one’s life (suicidal ideation), to formulating a specific plan (suicide planning), to acting on this plan with either a non-fatal outcome (suicide attempt) or a fatal outcome (suicide) (Nock, Borges, Bromet, Cha, et al., 2008; O’Connor & Kirtley, 2018). Suicidal trajectories can be non-linear (Bryan et al., 2020). Individuals may oscillate between ideation and planning (O’Connor & Kirtley, 2018). Suicide may also occur in isolation as an impulsive act without preceding stages (Bryan et al., 2020). Nonetheless, progression from ideation to planning, and particularly a history of suicide attempts, is associated with an increased risk of future suicidal behavior (de la Torre-Luque et al., 2023; Demesmaeker et al., 2022; Nock, Borges, Bromet, Cha, et al., 2008; O’Connor & Kirtley, 2018; Skrivankova et al., 2025).

Suicidal ideation, suicide planning, and suicide attempts, hereafter referred to as suicidal thoughts and behaviours (STBs), are common in South Africa. The most recent nationally representative survey (2002-2004) estimated a lifetime prevalence of 9.1% for suicide ideation, 3.8% for suicide planning, and 2.9% for suicide attempts (Joe et al., 2008). Suicide attempts were about twice as frequent in females (3.8%) than in males (1.8%) (Joe et al., 2008). Higher prevalence estimates have been identified in adolescents and among university students (Bantjes et al., 2023; Casale et al., 2019).

From a public health perspective, suicide and STBs arise from the interaction of structural and societal determinants, contextual stressors, and individual factors (Pirkis et al., 2024). Structural determinants include public and social policies, legislative and regulatory frameworks, access to health care, cultural norms, and social determinants, including exposure to violence (Pirkis et al., 2024; World Health Organization, 2014). Contextual stressors include stressful life events (O’Connor & Nock, 2014), and poverty (Iemmi et al., 2016). Individual factors include sociodemographic characteristics (age, sex, education, employment) (Favril et al., 2023; Li et al., 2011) and psychological and clinical conditions, most prominently mental disorders (Favril et al., 2023; Moitra et al., 2021), alcohol and substance use disorders (Athey et al., 2025), and clinical conditions (Favril et al., 2023), such as HIV (Haas et al., 2025). In addition, there are also predisposing factors, including personality traits (Bruno et al., 2023; O’Connor & Nock, 2014), genetic (DiBlasi et al., 2021), and neurobiological factors (van Heeringen & Mann, 2014).

Based on psychological autopsy studies reporting psychiatric disorders in more than 90% of people who died by suicide (Arsenault-Lapierre et al., 2004; Bertolote & Fleischmann, 2002), suicidal behaviour has historically been framed as a mental health problem, and research has focused on mental disorders as the key contributing factor (Pirkis et al., 2024). However, increasing evidence and contemporary theoretical models suggest a broader aetiology of STBs in which structural and social determinants of health also play an important role, both directly and indirectly, by increasing the risk of mental health conditions (Knipe et al., 2022; O’Connor & Kirtley, 2018; Pirkis et al., 2024). This is particularly relevant in LMICs, where mental disorders are estimated to be present in 58% of suicide deaths and 45% of non-fatal suicidal behaviour, suggesting a less dominant role than in high-income countries and highlighting the potential importance of contextual factors such as poverty, unemployment, economic insecurity, violence, and social disadvantage (Iemmi et al., 2016; Knipe et al., 2019). In these settings, social determinants may contribute to STBs through pathways related to chronic stress, exposure to violence, and adverse childhood experiences, and by limiting access to mental health care. Mental disorders, therefore, remain important, but they do not fully explain the burden of STBs, particularly in LMIC contexts.

We conducted a cross-sectional study of mental health and STBs in peri-urban communities in Cape Town, South Africa, among adults with and without HIV. These settings are characterised by persistent structural inequities, high unemployment, low incomes, limited formal housing, high levels of violence, trauma, and substance use, and a high burden of HIV and tuberculosis, creating a complex risk environment for adverse mental health outcomes and STBs. Few studies have systematically quantified the contributions of various psychosocial and structural determinants of STBs among people living with HIV (PWH), and evidence from South Africa remains scares. We aimed to estimate the prevalence of STBs in these communities to examine associations with socioeconomic factors (age, sex, marital status, education, employment), structural factors (violence exposure), perceived stress, and clinical factors (mental disorders, including substance use disorders, and HIV).

## Methods

### Study design and participants

We conducted a cross-sectional study at three health facilities in the Cape Town metropolitan area, South Africa, from March 2023 to September 2024. Recruitment followed a quota-based sampling frame to achieve equal numbers by sex (male/female) and age (18–29, 30–44, ≥45 years), with a target enrolment of two-thirds PWH and one-third HIV-negative participants. This design enabled comparison of mental health and suicidal behaviours between PWH and HIV-negative participants of similar age and sex. Recruitment took place during outpatient clinic visits. Most PWH were recruited from antiretroviral therapy (ART) clinics. HIV-negative participants were mainly recruited from general medical outpatient services or family planning clinics. Participants were either attending routine follow-up or acute care services at the time of recruitment. Eligible individuals were 18 years or older, spoke English, isiXhosa or Afrikaans, and provided informed consent. Research assistants administered mental health screening tools and structured questionnaires to collect sociodemographic, behavioural and medical information. They received training led by the project manager, a medical doctor and mental health researcher, on standardised administration of all study instruments, followed by supervised role-play. The project manager observed each research assistant’s initial participant interviews and conducted periodic quality assurance checks throughout data collection to ensure consistent administration across sites. Trained research nurses subsequently conducted a structured diagnostic interview. Nurses received a full-day training led by a psychiatrist experienced with the instrument, covering standardised administration and scoring, followed by supervised role-plays. The project manager observed each nurse’s first two diagnostic interviews and periodically reviewed completed forms. Nurses were blinded to the mental health screening results and could consult the project manager or psychiatrist regarding specific cases throughout the study. Nurses and research assistants conducted interviews in Afrikaans, isiXhosa, or English, depending on participants’ preferred language. Individuals with unknown HIV status underwent HIV testing.

### Setting

Participants were recruited from two public-sector primary care clinics in peri-urban communities on the Cape Flats (Facilities 1 and 2) and from a tertiary referral hospital in the Cape Town metropolitan area (Facility 3). The two peri-urban primary care clinics serve predominantly low-income communities established between the 1950s and 1970s by South Africa’s apartheid government to resettle families forcibly removed from other parts of Cape Town under the Group Areas Act. These removals displaced many families from extended family networks and established communities. In subsequent decades, the displaced communities experienced socioeconomic decline, high unemployment (particularly among youth), poverty, increased gang violence, drug use, and the growth of informal settlements, characterised by inadequate housing infrastructure (e.g., limited piped water and flush toilets) and limited access to mental health care (Ruffieux et al., 2021; Statistics South Africa (Stats SA), 2012). These enduring conditions contribute to increased levels of interpersonal, gender-based, and gang-related violence, although their intensity varies across communities. The tertiary hospital serves a diverse catchment population of more than 3 million people. Recruitment at Facility 3 ceased in June 2023 due to logistical challenges of recruiting participants from this facility.

### Outcomes

Outcomes were STBs in the past 30-days (suicidal ideation, suicide plan, suicide attempt) and lifetime suicide attempt, assessed using the MINI International Neuropsychiatric Interview version 6.0 (Sheehan et al., 1998). The MINI suicidality module includes 13 yes/no items referring to STBs in the past 30-days. We defined *suicidal ideation* as endorsement of at least one of the following: thinking one would be better off dead, wishing to be dead, thinking about hurting or injuring oneself, or thinking about killing oneself. A *suicide plan* was defined as endorsement of at least one of the following: having a method in mind, having a plan in mind, taking active steps to prepare a suicide attempt but being interrupted, or taking active steps to prepare to kill oneself without initiating the attempt. A *suicide attempt* was defined as endorsement of at least one of the following: starting an attempt but aborting, starting an attempt that was interrupted by someone else, or attempting suicide. In addition, we constructed a hierarchical four-level categorical variable for past 30-day STBs (none, ideation, plan, attempt), assigning each participant to the highest level endorsed. Lifetime suicide attempt was measured with the MINI item asking whether the participant had ever attempted suicide in their lifetime (yes/no).

### Exposures

Sociodemographic characteristics were collected using study-specific questionnaires, including age, sex (male or female), self-identified population group (Black African, South African Mixed Ancestry, Indian/Asian, White, or other/unspecified), employment in the past 12 months (none, employed in the past 12 months but not currently, or currently employed), educational attainment (no formal education, primary education, or secondary/tertiary), and marital status (single, married/living together, or widowed/separated/divorced).

HIV status was obtained from the clinic records. For participants not known to be living with HIV and without a documented test in the past three months, HIV testing was conducted according to the South African public sector algorithm. This algorithm uses a serial two-test approach with rapid tests, beginning with a high-sensitivity screening test, the Abon HIV rapid diagnostic test, with non-reactive results classified as HIV-negative, followed by a confirmatory test using a different assay (First Response HIV) for any reactive screening result. Individuals with two reactive results were diagnosed with HIV and referred for HIV care and treatment.

Current mental disorders were assessed using the MINI, version 6.0 (Sheehan et al., 1998), based on the DSM-IV diagnostic criteria. Assessed conditions included alcohol use disorder (including abuse or dependence), substance use disorders other than alcohol-related (abuse or dependence), psychotic disorders, bipolar and related disorders (mania or hypomania and mood episodes with psychotic features), major depressive episode, generalised anxiety disorder, post-traumatic stress disorder, and other anxiety disorders (panic disorder, agoraphobia, and social anxiety disorder).

Perceived stress in the past month was assessed at enrolment using the Perceived Stress Scale 4 (PSS-4), categorized as low (0-5), moderate (6-10), and high (11-16) (Cohen et al., 1983). Violence exposure was assessed with a study-specific questionnaire comprising two items asking whether the participant had ever experienced an act of violence in the community or violence in the family.

### Statistical analysis

Descriptive statistics were used to summarise sociodemographic, behavioural, and health-related characteristics by recruitment facility and the presence of past 30-day suicidal ideation. We estimated the prevalence of past 30-day suicidal ideation, suicide plan, suicide attempt, and lifetime suicide attempt by facility, with 95% logit-transformed binomial confidence intervals.

In the primary analysis, we used logistic regression to estimate unadjusted and adjusted odds ratios (ORs) for factors associated with past 30-day suicidal ideation. We fitted seven multivariable models. Model 1 adjusted for age group (18–29, 30–44, ≥45 years), sex, HIV status, and population group. Model 2 added sociodemographic factors (employment, education, marital status) and violence experience in the community and in the family to Model 1; Model 3 added the perceived stress score category based on the PSS-4 (low, moderate, high) to Model 1; Model 4 added all current mental disorders with prevalence >5% (major depressive episode, generalised anxiety disorder, post-traumatic stress disorder, other anxiety disorders, alcohol use disorder, substance use disorder) to Model 1; Model 5 included adjusted violence experience, and perceived stress for mental disorders in addition to Model 1 variables. Model 6 added a single predictor for the presence of any current mental disorder to Model 1. Model 7 included a predictor for the number of current mental disorders (none, 1, 2, 3, or more) to model 1. To facilitate interpretation on the absolute risk scale, we reported average marginal effects (AMEs) derived from the adjusted models. We estimated AMEs as average discrete changes in the predicted probability of suicidal ideation for each category relative to its reference group, averaged over the observed covariate distribution in the analytic sample. We present AMEs as percentage-point differences with 95% confidence intervals.

In a secondary analysis, we used ordinal logistic regression models to estimate unadjusted and adjusted ORs for factors associated with past 30-day STBs (none, ideation, plan, attempt). Ordinal logistic regression was selected to model this ordered categorical outcome, to account for the inherent ranking of severity in STBs. We fitted the same seven multivariable models as in the primary analysis.

We generated Venn/Euler diagrams, stratified by past 30-day suicidal ideation. Sets were defined from binary indicators: mental disorder (presence of any assessed current mental disorder), violence exposure (lifetime exposure within family or community), and high stress (PSS-4 ≥11). Areas represent approximate overlaps derived from Euler fits, and labels show participant counts.

Regression analyses used complete cases because missing data were rare. Analyses were done in Stata, version 18 (StataCorp LLC, College Station, TX, USA) and R, version 4.4.2 (R Foundation for Statistical Computing, Vienna, Austria).

### Ethical considerations

Ethical approval for the study was obtained from the University of Cape Town Human Research Ethics Committee (HREC, REF: 129/2022) and the Cantonal Ethics Committee of the Canton of Bern, Switzerland (KEK, REF: 150/14). Participants provided written informed consent. Following HREC guidance, we do not disclose facility names, exact locations, or detailed contextual information (e.g., population group and interview language) to avoid stigmatising communities. Interviewers received training to manage participant distress during the interviews. Individuals reporting active suicidality were referred to free mental health services on the day of the interview in line with the local standard suicide risk management protocols.

## Results

### Characteristics of the study population

Of 613 enrolled participants, 401 (65.4%) were recruited at Facility 1, 201 (32.8%) at Facility 2, and 11 (1.8%) at Facility 3. The target 2:1 recruitment ratio by HIV status was achieved (PWH, 65.4%), while females (63.9%) were slightly over-recruited relative to the 1:1 sex target. The median age of participants was 39 years (IQR 27–48) (Table 1).

**Table 1:**
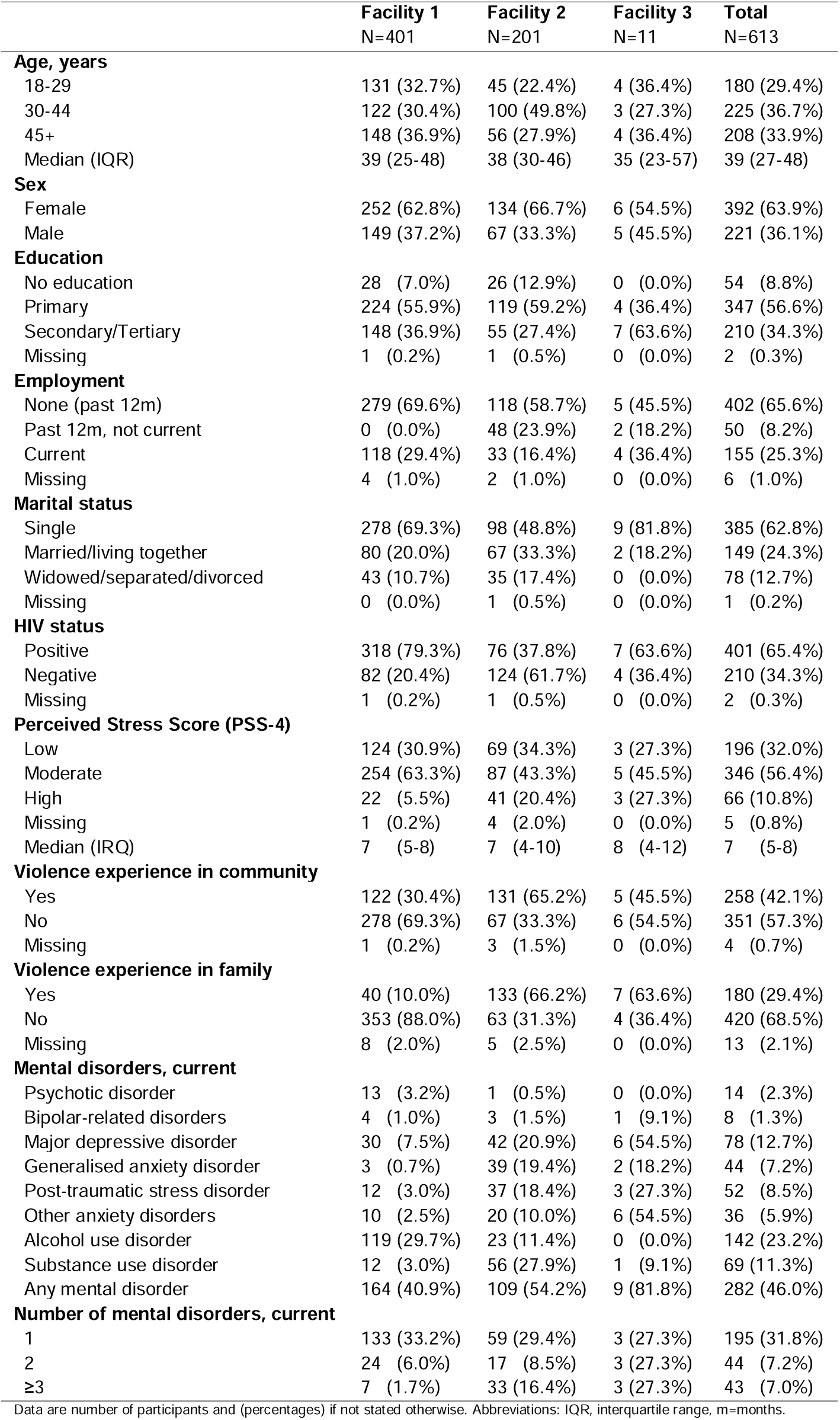
Characteristics of the study population by facility.

|  | Facility 1<br>N=401 | Facility 2<br>N=201 | Facility 3<br>N=11 | Total<br>N=613 |
| --- | --- | --- | --- | --- |
| **Age, years** |  |  |  |  |
| 18-29 | 131 (32.7%) | 45 (22.4%) | 4 (36.4%) | 180 (29.4%) |
| 30-44 | 122 (30.4%) | 100 (49.8%) | 3 (27.3%) | 225 (36.7%) |
| 45+ | 148 (36.9%) | 56 (27.9%) | 4 (36.4%) | 208 (33.9%) |
| Median (IQR) | 39 (25-48) | 38 (30-46) | 35 (23-57) | 39 (27-48) |
| **Sex** |  |  |  |  |
| Female | 252 (62.8%) | 134 (66.7%) | 6 (54.5%) | 392 (63.9%) |
| Male | 149 (37.2%) | 67 (33.3%) | 5 (45.5%) | 221 (36.1%) |
| **Education** |  |  |  |  |
| No education | 28 (7.0%) | 26 (12.9%) | 0 (0.0%) | 54 (8.8%) |
| Primary | 224 (55.9%) | 119 (59.2%) | 4 (36.4%) | 347 (56.6%) |
| Secondary/Tertiary | 148 (36.9%) | 55 (27.4%) | 7 (63.6%) | 210 (34.3%) |
| Missing | 1 (0.2%) | 1 (0.5%) | 0 (0.0%) | 2 (0.3%) |
| **Employment** |  |  |  |  |
| None (past 12m) | 279 (69.6%) | 118 (58.7%) | 5 (45.5%) | 402 (65.6%) |
| Past 12m, not current | 0 (0.0%) | 48 (23.9%) | 2 (18.2%) | 50 (8.2%) |
| Current | 118 (29.4%) | 33 (16.4%) | 4 (36.4%) | 155 (25.3%) |
| Missing | 4 (1.0%) | 2 (1.0%) | 0 (0.0%) | 6 (1.0%) |
| **Marital status** |  |  |  |  |
| Single | 278 (69.3%) | 98 (48.8%) | 9 (81.8%) | 385 (62.8%) |
| Married/living together | 80 (20.0%) | 67 (33.3%) | 2 (18.2%) | 149 (24.3%) |
| Widowed/separated/divorced | 43 (10.7%) | 35 (17.4%) | 0 (0.0%) | 78 (12.7%) |
| Missing | 0 (0.0%) | 1 (0.5%) | 0 (0.0%) | 1 (0.2%) |
| **HIV status** |  |  |  |  |
| Positive | 318 (79.3%) | 76 (37.8%) | 7 (63.6%) | 401 (65.4%) |
| Negative | 82 (20.4%) | 124 (61.7%) | 4 (36.4%) | 210 (34.3%) |
| Missing | 1 (0.2%) | 1 (0.5%) | 0 (0.0%) | 2 (0.3%) |
| **Perceived Stress Score (PSS-4)** |  |  |  |  |
| Low | 124 (30.9%) | 69 (34.3%) | 3 (27.3%) | 196 (32.0%) |
| Moderate | 254 (63.3%) | 87 (43.3%) | 5 (45.5%) | 346 (56.4%) |
| High | 22 (5.5%) | 41 (20.4%) | 3 (27.3%) | 66 (10.8%) |
| Missing | 1 (0.2%) | 4 (2.0%) | 0 (0.0%) | 5 (0.8%) |
| Median (IQR) | 7 (5-8) | 7 (4-10) | 8 (4-12) | 7 (5-8) |
| **Violence experience in community** |  |  |  |  |
| Yes | 122 (30.4%) | 131 (65.2%) | 5 (45.5%) | 258 (42.1%) |
| No | 278 (69.3%) | 67 (33.3%) | 6 (54.5%) | 351 (57.3%) |
| Missing | 1 (0.2%) | 3 (1.5%) | 0 (0.0%) | 4 (0.7%) |
| **Violence experience in family** |  |  |  |  |
| Yes | 40 (10.0%) | 133 (66.2%) | 7 (63.6%) | 180 (29.4%) |
| No | 353 (88.0%) | 63 (31.3%) | 4 (36.4%) | 420 (68.5%) |
| Missing | 8 (2.0%) | 5 (2.5%) | 0 (0.0%) | 13 (2.1%) |
| **Mental disorders, current** |  |  |  |  |
| Psychotic disorder | 13 (3.2%) | 1 (0.5%) | 0 (0.0%) | 14 (2.3%) |
| Bipolar-related disorders | 4 (1.0%) | 3 (1.5%) | 1 (9.1%) | 8 (1.3%) |
| Major depressive disorder | 30 (7.5%) | 42 (20.9%) | 6 (54.5%) | 78 (12.7%) |
| Generalised anxiety disorder | 3 (0.7%) | 39 (19.4%) | 2 (18.2%) | 44 (7.2%) |
| Post-traumatic stress disorder | 12 (3.0%) | 37 (18.4%) | 3 (27.3%) | 52 (8.5%) |
| Other anxiety disorders | 10 (2.5%) | 20 (10.0%) | 6 (54.5%) | 36 (5.9%) |
| Alcohol use disorder | 119 (29.7%) | 23 (11.4%) | 0 (0.0%) | 142 (23.2%) |
| Substance use disorder | 12 (3.0%) | 56 (27.9%) | 1 (9.1%) | 69 (11.3%) |
| Any mental disorder | 164 (40.9%) | 109 (54.2%) | 9 (81.8%) | 282 (46.0%) |
| **Number of mental disorders, current** |  |  |  |  |
| 1 | 133 (33.2%) | 59 (29.4%) | 3 (27.3%) | 195 (31.8%) |
| 2 | 24 (6.0%) | 17 (8.5%) | 3 (27.3%) | 44 (7.2%) |
| ≥3 | 7 (1.7%) | 33 (16.4%) | 3 (27.3%) | 43 (7.0%) |

Data are number of participants and (percentages) if not stated otherwise. Abbreviations: IQR, interquartile range, m=months.

Most participants had completed primary education (56.6%), were single (62.8%), and had not been employed in the past 12 months (65.6%). Perceived stress was moderate in 56.4% and high in 10.8%. Overall, 42.1% reported ever experiencing an act of violence in their community, 29.4% reported ever experiencing violence in their family, and 22% ever experiencing both violence in the community and violence in their family, with higher prevalences at Facilities 2 and 3 than at Facility 1 (Table 1).

The prevalence of current mental disorders was high. Overall, alcohol use disorder was most common (23.2%), followed by major depressive disorder (12.7%), substance use disorder (11.3%), post-traumatic stress disorder (8.5%), and generalised anxiety disorder (7.2%). Patterns varied by facility: Facility 1 had a high prevalence of alcohol use disorder with comparatively low prevalences of other disorders. Facility 2 showed higher prevalences of major depressive disorder, anxiety disorders, and substance use disorders, with a lower prevalence of alcohol use disorder. Facility 3 had very high prevalences of major depressive disorder and post-traumatic stress disorder, although numbers were small (n=11) (Table 1).

### Prevalence of suicidal thoughts and behaviours (STBs)

The prevalence of STBs by sex, HIV status, and facility is shown in Table 2. Overall, 14.0% of participants reported past 30-day suicidal ideation (95% CI 11.5-17.0). Among participants with ideation, 22.1% (95% CI 14.5-32.2) reported a suicide plan, and 16.3% (95% CI 9.8-25.8) reported a suicide attempt in the past 30 days. Facilities 1 and 2 had similar prevalence of ideation, but plans and attempts among those with ideation were more common at Facility 2 than at Facility 1. Lifetime suicide attempt prevalence was 22.2% (95% CI 19.1-30.9) overall and was slightly higher at Facilities 2 and 3.

**Table 2:** Prevalence of suicidal thoughts and behaviors by sex, HIV status and facility.

|  | Facility 1 |  |  | Facility 2 |  |  | Facility 3 |  |  | Total |  |  |
| --- | --- | --- | --- | --- | --- | --- | --- | --- | --- | --- | --- | --- |
|  | N | Events | Prevalence (95% CI) | N | Events | Prevalence (95% CI) | N | Events | Prevalence (95% CI) | N | Events | Prevalence (95% CI) |
| **Overall** |  |  |  |  |  |  |  |  |  |  |  |  |
| Ideation, past 30 days | 401 | 52 | 13.0% (10.0-16.6) | 201 | 30 | 14.9% (10.6-20.6) | 11 | 4 | 36.4% (12.4-69.8) | 613 | 86 | 14.0% (11.5-17.0) |
| Plan, past 30 days | 52 | 4 | 7.7% (2.8-19.2) | 30 | 14 | 46.7% (29.3-64.9) | 4 | 1 | 25.0% (0.8-92.9) | 86 | 19 | 22.1% (14.5-32.2) |
| Attempt, past 30 days | 52 | 4 | 7.7% (2.8-19.2) | 30 | 10 | 33.3% (18.5-52.5) | 4 | 0 | 0.0% (0.0-60.2) | 86 | 14 | 16.3% (9.8-25.8) |
| Attempt, lifetime | 401 | 81 | 20.2% (16.5-24.4) | 201 | 52 | 25.9% (20.3-32.4) | 11 | 3 | 27.3% (7.7-62.9) | 613 | 136 | 22.2% (19.1-25.7) |
| **Female** |  |  |  |  |  |  |  |  |  |  |  |  |
| Ideation, past 30 days | 252 | 40 | 15.9% (11.8-20.9) | 134 | 24 | 17.9% (12.3-25.4) | 6 | 2 | 33.3% (5.1-82.2) | 392 | 66 | 16.8% (13.4-20.9) |
| Plan, past 30 days | 40 | 4 | 10.0% (3.7-24.4) | 24 | 11 | 45.8% (26.6-66.4) | 2 | 1 | 50.0% (0.0-100.0) | 66 | 16 | 24.2% (15.3-36.2) |
| Attempt, past 30 days | 40 | 3 | 7.5% (2.4-21.4) | 24 | 8 | 33.3% (17.0-55.0) | 2 | 0 | 0.0% (0.0-84.2) | 66 | 11 | 16.7% (9.4-27.9) |
| Attempt, lifetime | 252 | 63 | 25.0% (20.0-30.7) | 134 | 40 | 29.9% (22.7-38.2) | 6 | 0 | 0.0% (0.0-77.6) | 392 | 103 | 26.3% (22.1-30.9) |
| **Male** |  |  |  |  |  |  |  |  |  |  |  |  |
| Ideation, past 30 days | 149 | 12 | 8.1% (4.6-13.7) | 67 | 6 | 9.0% (4.0-18.8) | 5 | 2 | 40.0% (5.0-89.4) | 221 | 20 | 9.0% (5.9-13.6) |
| Plan, past 30 days | 12 | 0 | 0.0% (0.0-26.5) | 6 | 3 | 50.0% (10.9-89.1) | 2 | 0 | 0.0% (0.0-84.2) | 20 | 3 | 15.0% (4.5-39.6) |
| Attempt, past 30 days | 12 | 1 | 8.3% (0.9-47.5) | 6 | 2 | 33.3% (5.1-82.2) | 2 | 0 | 0.0% (0.0-84.2) | 20 | 3 | 15.0% (4.5-39.6) |
| Attempt, lifetime | 149 | 18 | 12.1% (7.7-18.4) | 67 | 12 | 17.9% (10.4-29.2) | 5 | 3 | 60.0% (10.6-95.0) | 221 | 33 | 14.9% (10.8-20.3) |
| **HIV-positive** |  |  |  |  |  |  |  |  |  |  |  |  |
| Ideation, past 30 days | 318 | 38 | 11.9% (8.8-16.0) | 76 | 15 | 19.7% (12.2-30.4) | 7 | 3 | 42.9% (10.4-82.9) | 401 | 56 | 14.0% (10.9-17.7) |
| Plan, past 30 days | 38 | 3 | 7.9% (2.5-22.5) | 15 | 8 | 53.3% (27.4-77.6) | 3 | 0 | 0.0% (0.0-70.8) | 56 | 11 | 19.6% (11.1-32.4) |
| Attempt, past 30 days | 38 | 3 | 7.9% (2.5-22.5) | 15 | 8 | 53.3% (27.4-77.6) | 3 | 0 | 0.0% (0.0-70.8) | 56 | 11 | 19.6% (11.1-32.4) |
| Attempt, lifetime | 318 | 67 | 21.1% (16.9-25.9) | 76 | 24 | 31.6% (22.0-43.0) | 7 | 3 | 42.9% (10.4-82.9) | 401 | 94 | 23.4% (19.5-27.9) |
| **HIV-negative** |  |  |  |  |  |  |  |  |  |  |  |  |
| Ideation, past 30 days | 82 | 14 | 17.1% (10.3-27.0) | 124 | 15 | 12.1% (7.4-19.2) | 4 | 1 | 25.0% (0.8-92.9) | 210 | 30 | 14.3% (10.2-19.7) |
| Plan, past 30 days | 14 | 1 | 7.1% (0.8-42.0) | 15 | 6 | 40.0% (17.7-67.4) | 1 | 1 | 100% (2.5-100) | 30 | 8 | 26.7% (13.5-45.8) |
| Attempt, past 30 days | 14 | 1 | 7.1% (0.8-42.0) | 15 | 2 | 13.3% (2.9-44.0) | 1 | 0 | 0.0% (0.0-97.5%) | 30 | 3 | 10.0% (3.1-27.8) |
| Attempt, lifetime | 82 | 14 | 17.1% (10.3-27.0) | 124 | 28 | 22.6% (16.0-30.9) | 4 | 0 | 0.0% (0.0-60.2) | 210 | 42 | 20.0% (15.1-26.0) |

CI, confidence interval.

The prevalence of STBs was higher in females than in males. Past 30-day suicidal ideation was reported by 16.8% of females and 9.0% of males. Lifetime suicide attempt was reported by 26.3% of females and 14.9% of males. Prevalence was similar by HIV status: among PWH, 14.0% reported past 30-day ideation and 23.4% reported a lifetime suicide attempt, compared with 14.3% and 20.0% among HIV-negative participants (Table 2). The hierarchically coded four-level measure of past 30-day STBs (none, ideation, plan, attempt) is shown in Table S1.

### Prevalence of mental disorders among participants with ideation

Among the 86 participants with past 30-day suicidal ideation, 81.4% had a current mental disorder, most frequently major depressive disorder (45.3%), post-traumatic stress disorder (34.9%), and alcohol use disorder (32.6%) (Table 3).

**Table 3:**
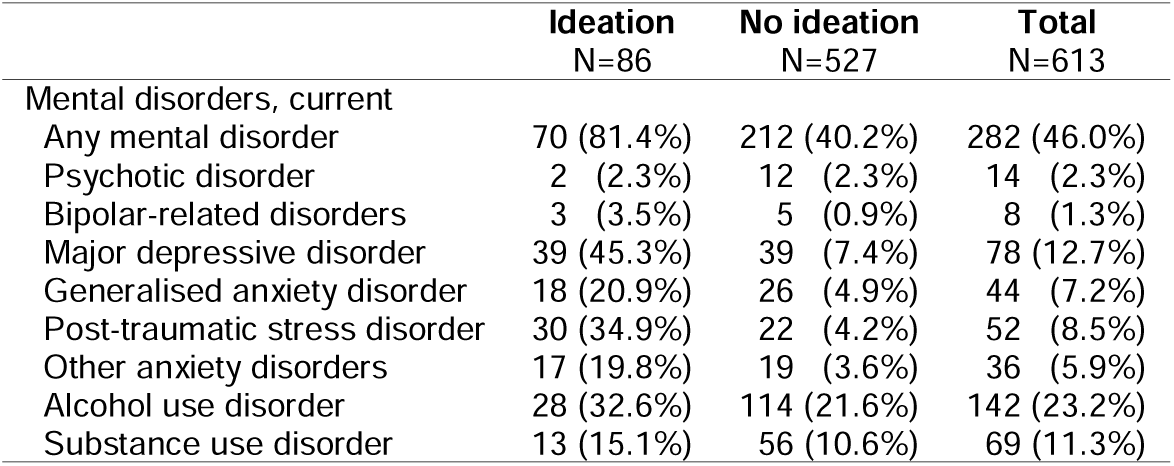
Prevalence of current mental disorders by past 30-day suicidal ideation.

### Co-occurrence of perceived stress, violence exposure, and mental disorders

The co-occurrence of high perceived stress, violence exposure, and current mental disorders in participants with and without past 30-day suicidal ideation is shown in Figure S1.

In participants with ideation, only a few participants did not have at least one of the three risk factors (5.8%), there was pronounced overlap between mental disorder, and violence exposure, and between mental disorders and high perceived stress.

### Factors associated with past 30-day suicidal ideation

#### HIV status

PWH (14.0%) and people without HIV (14.3%) reported a similar prevalence of suicidal ideation (Table 2). In Model 1, including HIV status, age group, sex, and population group, we found no evidence that HIV status was associated with past 30-day suicidal ideation (AME 2.01 percentage points, 95% CI −4.21 to 8.23) (Figure 1). We present the corresponding adjusted ORs in Figure S2.

**Figure 1:**
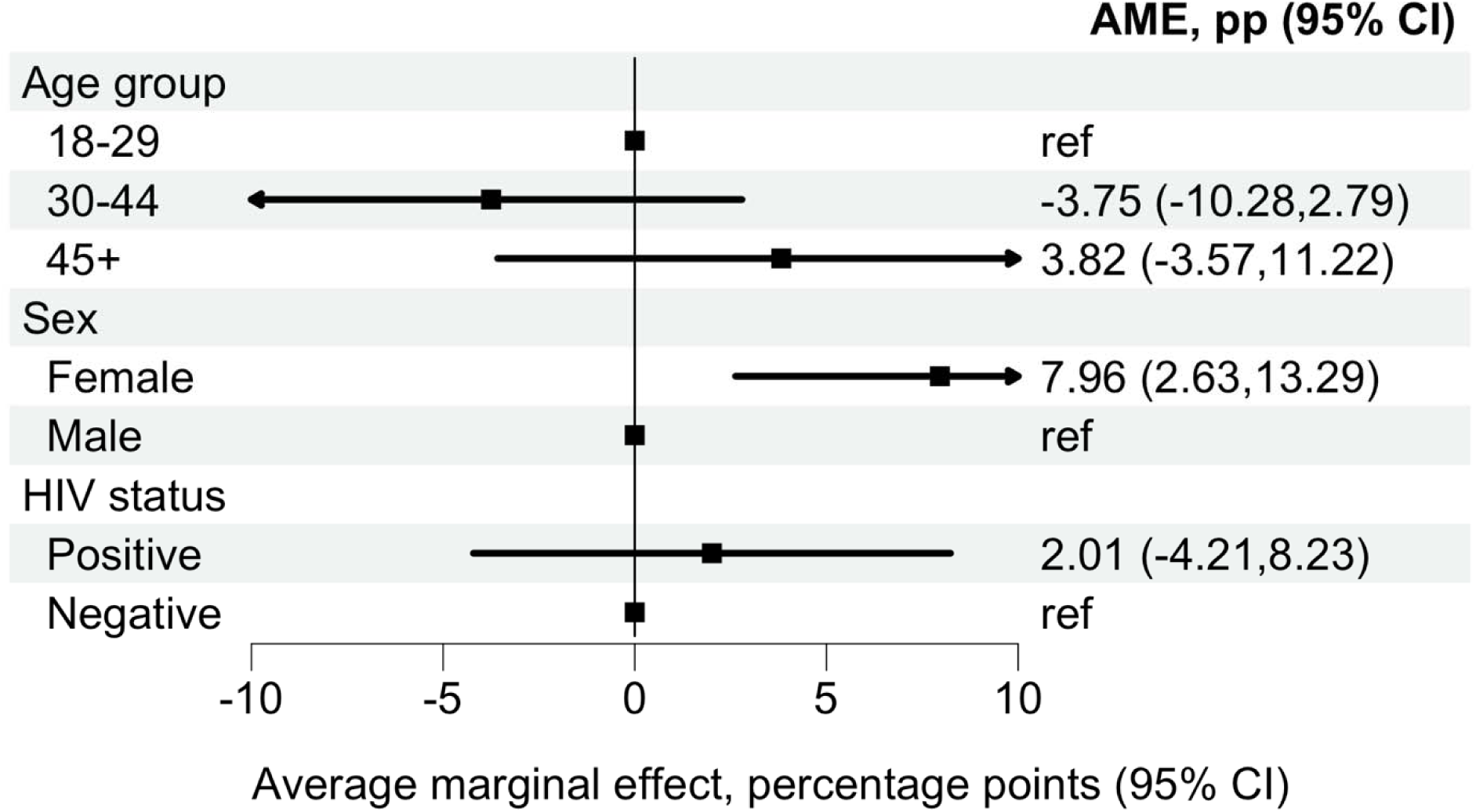
Average marginal effect of HIV status on past 30-day suicidal ideation. Average marginal effect (AME), expressed as a percentage-point difference, comparing people living with and without HIV from an adjusted logistic regression model for past 30-day suicidal ideation. The model adjusts for age group, sex, and population group. The AME represents the average discrete change in the predicted probability of suicidal ideation associated with HIV-positive status relative to HIV-negative status, averaged over the observed covariate distribution. Error bars indicate 95% confidence intervals. The corresponding adjusted odds ratio are presented in Figure S2.

#### Contextual factors (sociodemographic and violence)

In Model 2, which included sociodemographic characteristics, violence and HIV status, sex and violence exposure showed the clearest associations with past 30-day suicidal ideation. Female sex was associated with a higher predicted probability compared with male sex (AME 8.52 percentage points, 95% CI 3.08 to 13.96). Participants reporting violence in the community also had a higher predicted probability relative to those not reporting community violence (AME 7.52 percentage points, 95% CI 0.87 to 14.17). Reported family violence showed the largest estimated association, with a higher predicted probability of suicidal thoughts and behaviours (AME 14.09 percentage points, 95% CI 4.56 to 23.62). There was no evidence of association with age group, employment, educational attainment, marital status, or HIV status (Figure 2, Figure S3).

**Figure 2:**
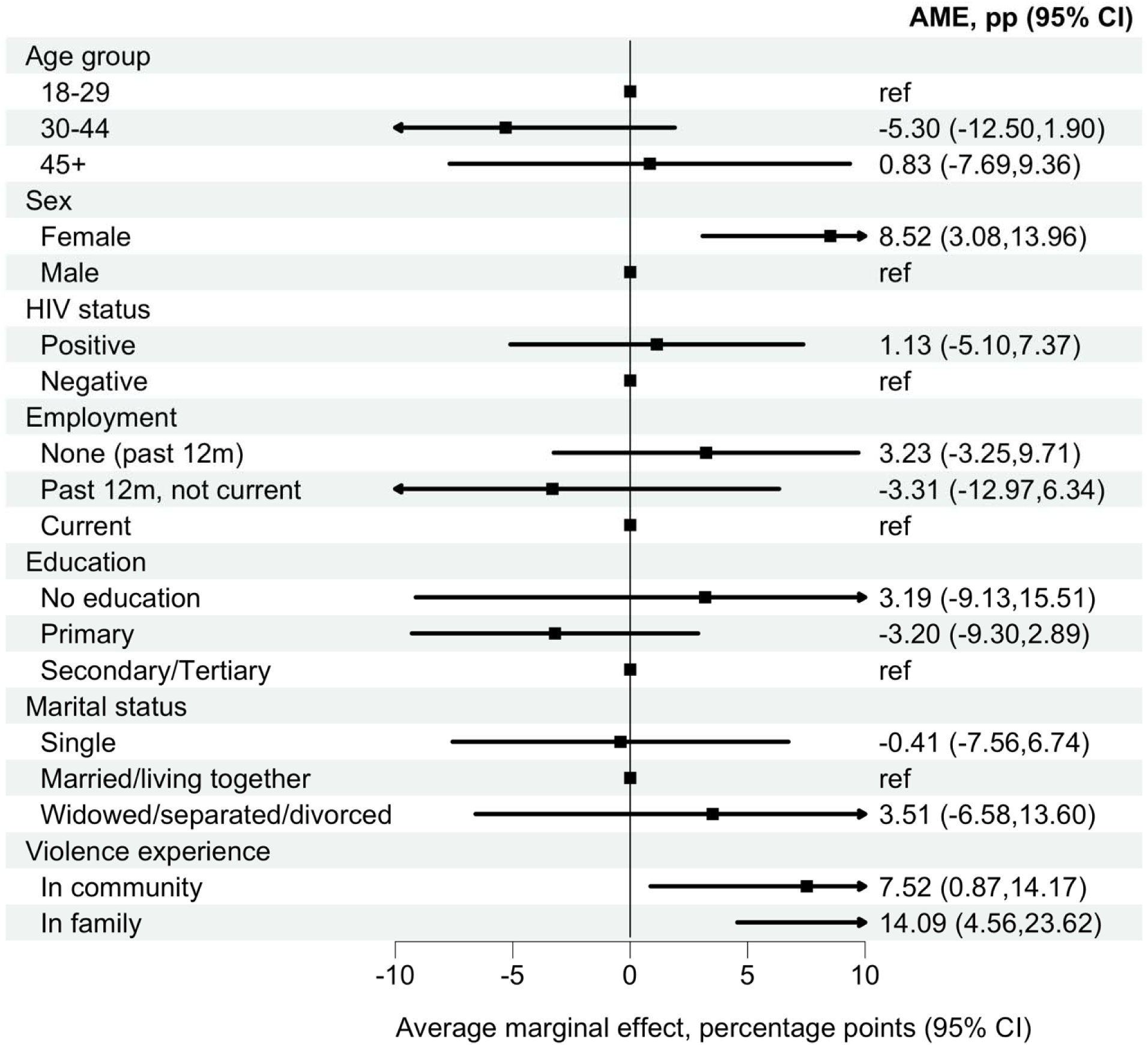
Average marginal effects for sociodemographic factors and violence exposure associated with past 30-day suicidal ideation. Average marginal effects (AMEs), expressed as percentage-point differences, for associations of sociodemographic characteristics and violence exposure with past 30-day suicidal ideation. Estimates derive from an adjusted logistic regression model including age group, sex, HIV status, employment, education, marital status, and violence exposure. AMEs represent the average discrete change in the predicted probability of suicidal ideation for each category relative to its reference group, averaged over the observe covariate distribution. Error bars indicate 95% confidence intervals. The corresponding adjusted odds ratio are presented in Figure S3.

#### Perceived stress

In Model 3, adjusted for age group, sex, population group, and HIV status, perceived stress was strongly associated with past 30-day suicidal ideation. Compared with participants with low perceived stress (PSS-4), those with moderate perceived stress had a higher predicted probability of suicidal ideation (AME 7.76 percentage points, 95% CI 2.76 to 12.77).

Participants with high perceived stress showed a markedly higher predicted probability (AME 33.58 percentage points, 95% CI 20.67 to 46.50). (Figure 3, Figure S4)

**Figure 3:**
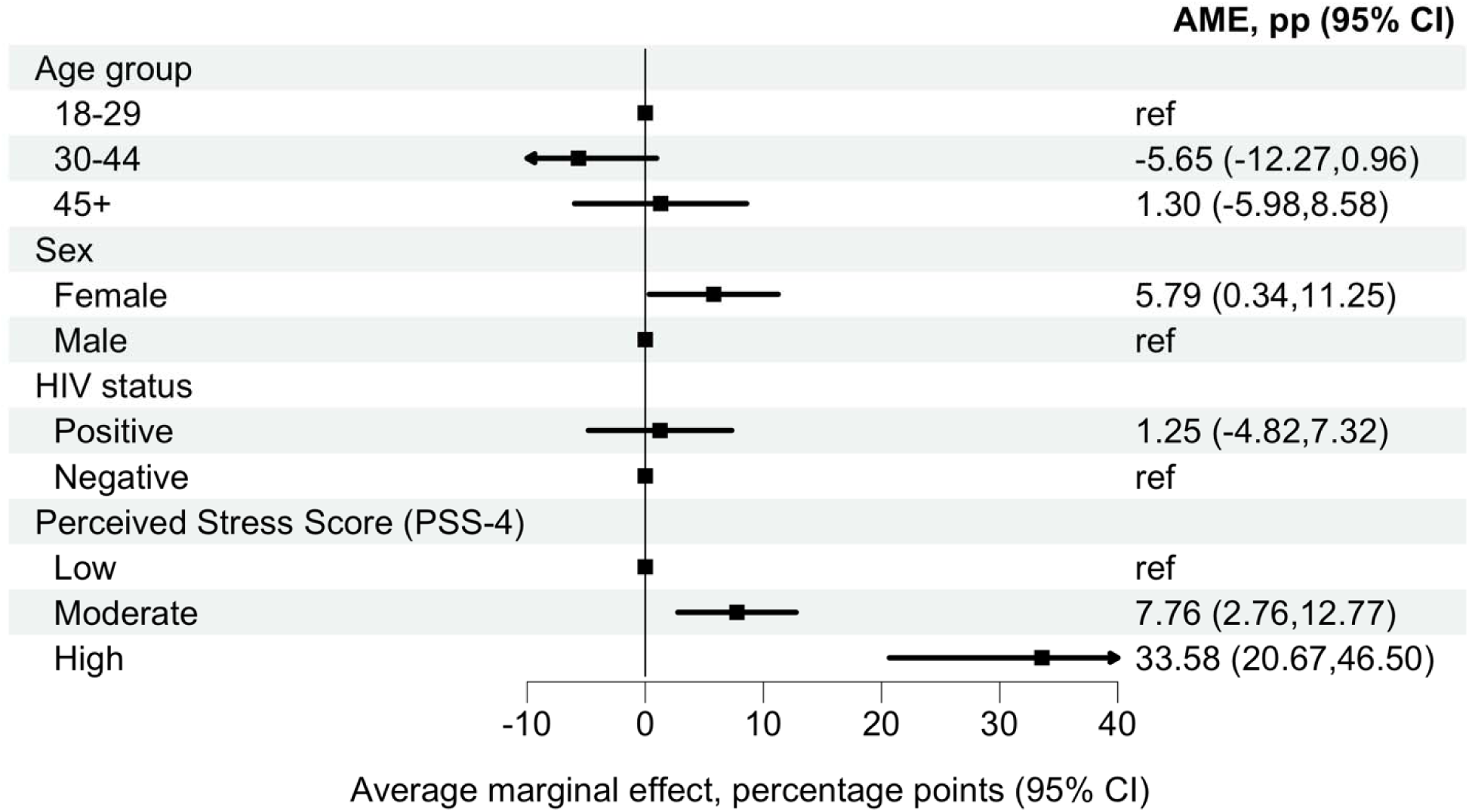
Average marginal effect for perceived stress with past 30-day suicidal ideation. Average marginal effects (AMEs), expressed as percentage-point differences, for associations of perceived stress (PSS-4 categories) with past 30-day suicidal ideation. Estimates derive from an adjusted logistic regression model including perceive stress category (low, moderate, high), age group, sex, and HIV status. AMEs represent the average discrete change in the predicte probability of suicidal ideation for each category relative to its reference group, averaged over the observed covariate distribution. Error bars indicate 95% confidence intervals. The corresponding adjusted odds ratio are presented in Figure S4.

#### Mental disorders

In Model 4, adjusted for age group, sex, population group, and HIV status, major depressive disorder (AME 27.13 percentage points, 95% CI 14.67 to 39.58) and post-traumatic stress disorder (AME 27.98 percentage points, 95% CI 12.12 to 43.84) showed the largest increases in the predicted probability of past 30-day suicidal ideation. Alcohol use disorder was also associated with a higher predicted probability (AME 8.34 percentage points, 95% CI 1.53 to 15.15). There was no evidence of associations with generalised anxiety disorder, other anxiety disorders, or substance use disorder (Figure 4, Figure S5). In a model adjusted for age group, sex, population group, and HIV status, the presence of any mental disorder was associated with a high predicted probability (AME 21.44 [95% CI 15.73, 27.15]) (Figures S6-S7). A dose-response increase was observed with a higher number of current mental disorders (1: AME 14.50 [95% CI 8.43, 20.56]; 2: AME 29.93 [15.94, 43.92]; ≥3: AME 49.43 [33.05, 65.80]) (Figures S8-S9).

**Figure 4:**
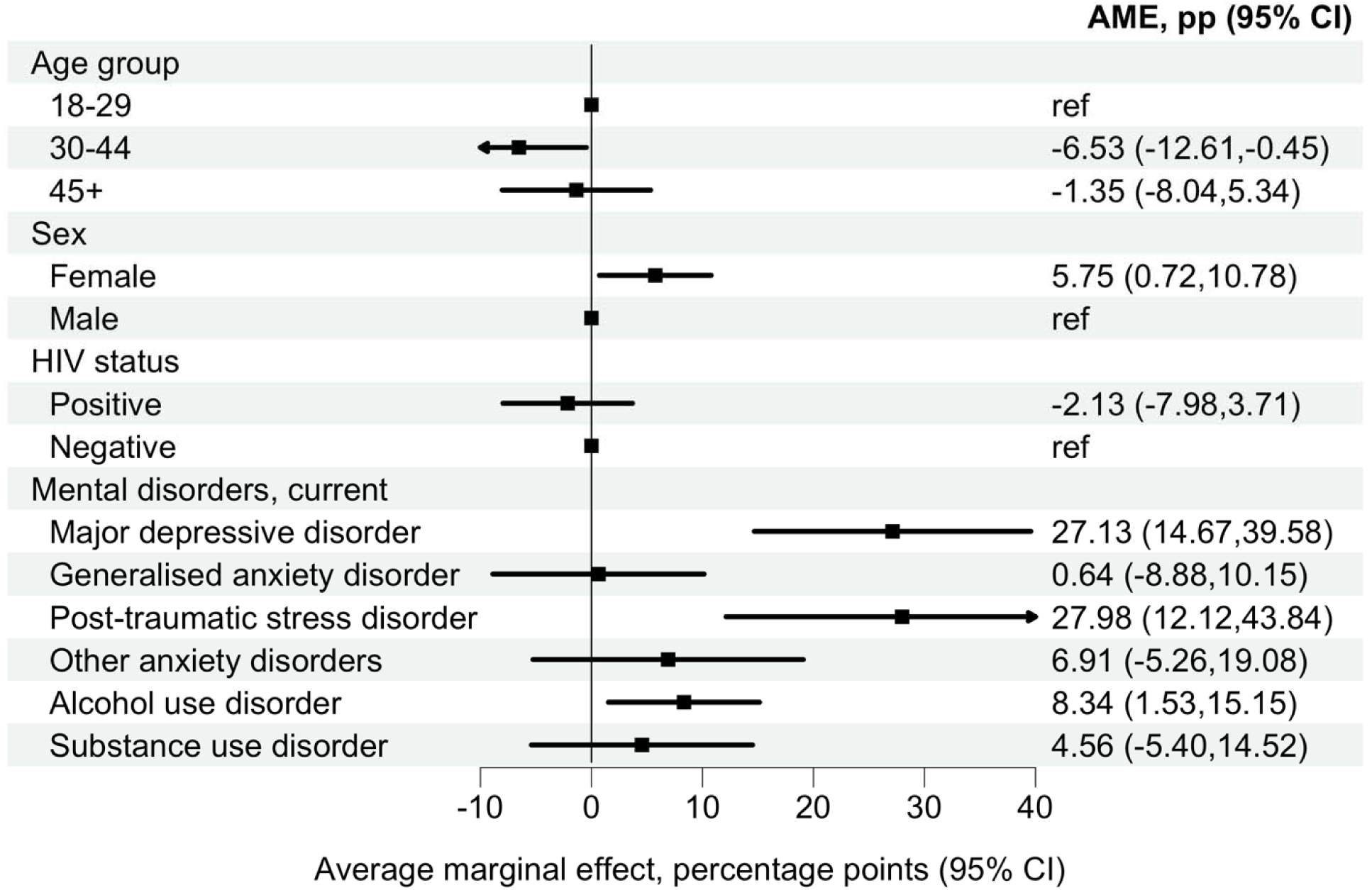
Average marginal effects for specific current mental disorders associated with past 30-day suicidal ideation. Average marginal effects (AMEs), expressed as percentage-point differences, for associations of current mental disorders with past 30-day suicidal ideation. Estimates derive from an adjusted logistic regression model including age group, sex, HIV status, and indicators for current mental disorders (major depressive disorder, generalised anxiety disorder, post-traumatic stress disorder, other anxiety disorders, alcohol use disorder, and substance use disorder). AMEs represent the average discrete change in th predicted probability of suicidal ideation associated with each disorder relative to the absence of that disorder, averaged over the observed covariate distribution. Error bars indicate 95% confidence intervals. The corresponding adjusted odds ratio are presented in Figure S5.

#### Violence experience and perceived stress adjusted for mental disorders

In Model 5, adjusted for age group, sex, population group, HIV status, perceived stress, violence exposure, and any current mental disorder, high versus low perceived stress remained associated with a higher predicted probability of past 30-day suicidal ideation (AME 21.64 percentage points, 95% CI 10.63 to 32.64). Violence exposure in the community also remained associated with a higher predicted probability (AME 6.22 percentage points, 95% CI 0.44 to 12.00). Violence exposure in the family was attenuated, with no clear evidence of association (AME 6.09 percentage points, 95% CI −1.41 to 13.60) (Figure 5, Figure S10).

**Figure 5:**
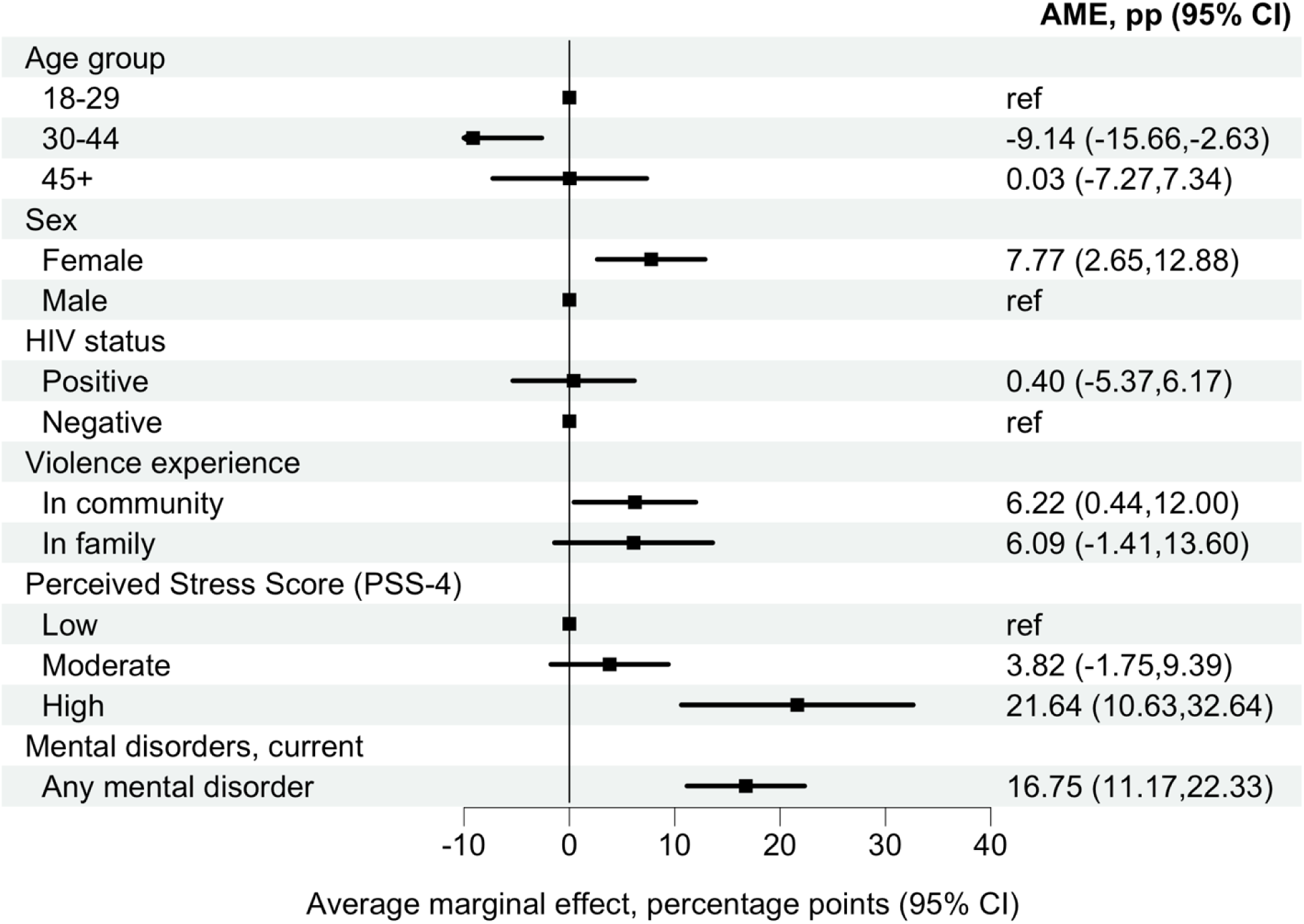
Average marginal effects for perceived stress and violence exposure associated with past 30-day suicidal ideation, adjusted for mental disorders. Average marginal effects (AMEs), expressed as percentage-point differences, for associations of perceived stress (PSS-4 categories) and violence exposure with past 30-day suicidal ideation. Estimates derive from an adjusted logistic regression model including perceived stress category and violence exposure, and adjusting for age group, sex, population group, HIV status, and the presence of any current mental disorder. AMEs represent the average discrete change in the predicted probability of suicidal ideation for each category relative to its reference group, averaged over the observed covariate distribution. Error bars indicate 95% confidence intervals.

### Factors associated with past 30-day suicidal behaviour

Odds ratios from ordinal logistic regression for the hierarchically coded STB outcome (none, ideation, plan, attempt) were very similar to those from the binary logistic regression models of suicidal ideation (Figure S2–Figure S5, Figure S7, Figure S9-Figure S10). Unadjusted ORs for all assessed associations are reported in Figure S11.

## Discussion

Past 30-day STBs (14.0%) and lifetime suicide attempts (22.2%) were highly prevalent in two socially deprived peri-urban communities in Cape Town. These estimates far exceed nationally representative figures from a household survey (2002–2004), which reported lifetime suicide attempts of 2.9% (Joe et al., 2008). This observed difference in prevalence likely reflects differences in the time period, sampling frame, and local context, as our study was conducted in two high-adversity urban communities rather than in a nationally representative sample. Past 30-day ideation was strongly associated with female sex, current mental disorders, higher levels of perceived stress, and exposure to violence. Associations with high perceived stress and community violence persisted after adjustment for mental disorders. There was no statistical evidence of association between past 30-day ideation and HIV status, marital status, education, or employment. Findings were unchanged when modelling past 30-day STBs as an ordinal outcome.

Our study aimed to clarify the relative importance of mental disorders and contextual factors in the aetiology of STB in South Africa by quantifying the associations of mental disorders, socioeconomic characteristics, perceived stress, and violence exposure with past 30-day suicidal ideation. High perceived stress was the strongest correlate of STB, exceeding associations observed for mental disorders. Mental disorders nonetheless remained highly prevalent among participants reporting ideation, affecting 80%, and they showed strong associations with past 30-day STB. Among specific conditions, major depressive disorder and post-traumatic stress disorder showed the largest increases in the predicted probability, each greater than a 27% increase. These findings are consistent with evidence from a recent study of 1.3 million South African medical insurance beneficiaries: 74% of individuals with a healthcare encounter for self-harm had a diagnosed mental disorder, and mental disorders were the strongest predictors of self-harm (Skrivankova et al., 2025). Associations between mental disorders and STBs observed in our study exceed estimates from other LMICs, where any mental disorder was associated with 3.6-fold higher odds of ideation, and mood or anxiety disorders with about threefold higher odds (Nock, Borges, Bromet, Alonso, et al., 2008).

Exposure to violence and perceived stress were independently associated with past 30-day STB after adjustment for mental disorders. This finding is consistent with earlier studies from South Africa reporting associations between interpersonal violence, sexual violence, and childhood and adulthood violence with suicidal ideation (Mkhwanazi et al., 2025; Sorsdahl et al., 2011). Our analysis extends this literature by showing that these contextual and psychosocial associations persist after adjustment for diagnosed mental disorders, suggesting that these factors may contribute to STBs independently of current mental health diagnoses. These findings should be interpreted within the broader structural context of peri-urban Cape Town, where spatial inequality, poverty, and limited access to services intersect with high levels of interpersonal and community violence. Such chronic social stressors may shape suicide risk beyond diagnosed mental disorders through cumulative stress and interpersonal conflict. Recent prospective research in South Africa has similarly identified informal housing and interpersonal conflict as predictors of subsequent suicide attempts, highlighting the role of contextual risk factors (Hutchison et al., 2025). Although our study did not include detailed measures of housing instability or social connectedness, these structural conditions likely form an important backdrop to the elevated prevalence of suicidal thoughts and behaviours observed. Longitudinal and mixed-methods studies are needed to clarify these pathways. Consistent with earlier evidence from South Africa, we found no evidence of association between other contextual factors, including employment, education, or marital status and suicidal ideation (Joe et al., 2008).

In this study, there was no evidence of association between living with HIV and past 30-day suicidal ideation (AME 2.01 percentage points, 95% CI −4.21 to 8.23; OR 1.19, 95% CI 0.69–2.04) or past 30-day STBs (OR 1.27, 95% CI 0.73–2.19). Among pregnant women in rural South Africa, the prevalence of ideation did not differ by HIV status (Rochat et al., 2013). By contrast, in Durban, a higher prevalence of ideation was observed 72 hours and 6 weeks after HIV testing among individuals who received a positive versus a negative result (Schlebusch & Govender, 2012). A large South African cohort study reported a 19% higher risk of healthcare presentation for intentional self-harm among PWH compared with HIV-negative individuals (Skrivankova et al., 2025). To the best of our knowledge, there are no African data on associations between HIV status and suicide mortality. Studies from Europe and North America consistently show higher suicide mortality in PWH compared with the general population, with a meta-analysis of 12 cohort studies indicating a fourfold higher risk in those living with HIV (Haas et al., 2025). However, results from these settings, where HIV primarily affects men who have sex with men and people who inject drugs, are not generalisable to South Africa’s generalised HIV epidemic. While findings remain inconsistent, HIV may be one of many contributing factors to suicide and STBs, rather than a causal factor. Furthermore, recent evidence suggests that PWH on antiretroviral therapy (ART) may have a similar risk profile for STBs as HIV-negative participants. Most of our participants living with HIV were probably established on ART, as they were recruited from ART clinics. However, we cannot confirm ART use because we did not collect data on treatment.

This study has several important strengths. We conducted this study in South African peri-urban communities characterised by high levels of socioeconomic deprivation, and structural and contextual stressors, creating an environment for poor mental and physical health outcomes and high rates of STBs. Evidence from such settings remains scarce. Our study helps address this gap, generating evidence to inform contextually relevant suicide prevention interventions addressing the needs of underserved and vulnerable populations. We collected a rich and comprehensive dataset. Trained and supervised mental health nurses used the MINI to assess STBs and a wide range of mental disorders. The use of a structured diagnostic interview with high sensitivity and specificity reduces misclassification of psychiatric diagnoses and provides nuanced data on a broad spectrum of STBs (Sheehan et al., 1998). In addition, we assessed socioeconomic, structural, psychosocial, and contextual factors, including exposure to violence and perceived stress. The breadth of this dataset allowed us to examine contextual and psychosocial correlates of STBs while adjusting for mental disorders.

Our results should be considered in the light of the following limitations. First, our cross-sectional study design is susceptible to prevalence-incidence (Neyman) bias (Hill, 2003). Individuals who died by suicide before data collection could not be included. Conditioning on survival can lead to an underestimation of STB prevalence and may bias associations for risk factors of suicide, typically towards the null. Because suicide mortality is a rare outcome, the magnitude of this bias is likely modest. Second, we could not assess associations for relevant low-prevalence disorders, such as bipolar disorder. Adequately assessing these associations would require very large samples. Third, we recruited participants in health facilities. Although 98% were enrolled in primary care clinics, STBs may be more prevalent among care-seeking populations than in the community. Fourth, while we generally used well-established, validated instruments to measure variables of interest, including the MINI neuropsychiatric interview and established mental health scales, our violence exposure question did not use a validated instrument, which may limit measurement precision. Fifth, our cross-sectional design limits our ability to determine the direction of causality between exposures and outcomes. For example, a bidirectional relationship between perceived stress and suicidal ideation is plausible. Perceived stress may contribute to suicidal ideation, but individuals experiencing suicidal ideation may also perceive their lives as more stressful. Several longitudinal studies suggest that perceived stress precedes the onset or worsening of suicidal ideation (Choi et al., 2022; Themelis et al., 2023)but we cannot exclude reverse causation in this study.

## Conclusion

This study underscores the importance of high perceived stress and violence exposure, in addition to mental disorders, as correlates of STBs in urban peri-urbans in South Africa, while challenging the assumption about the role of HIV as a key driver. Effective suicide prevention in these communities may require multilevel interventions that reduce psychosocial stress and violence, strengthen adaptive coping and stress management skills, and scale up access to quality mental health services.

## Data Availability

Data were obtained from the International epidemiology Databases to Evaluate AIDS-Southern Africa (IeDEA-SA). Data cannot be made available online because of legal and ethical restrictions. To request data, readers may contact IeDEA-SA for consideration by filling out the online form available at https://www.iedea-sa.org/contact-us/.

## Appendix

**Table S1:** Prevalence of past 30-day suicidal thoughts and behaviours, hierarchically coded, by facility.

|  | Facility 1<br>N=401 | Facility 2<br>N=201 | Facility 3<br>N=11 | Total<br>N=613 |
| --- | --- | --- | --- | --- |
| Past 30-day suicidal thoughts and behaviours |  |  |  |  |
| None | 349 (87.0%) | 171 (85.1%) | 7 (63.6%) | 527 (86.0%) |
| Suicidal ideation | 45 (11.2%) | 15 (7.5%) | 3 (27.3%) | 63 (10.3%) |
| Suicidal plan | 3 (0.7%) | 5 (2.5%) | 1 (9.1%) | 9 (1.5%) |
| Suicide attempt | 4 (1.0%) | 10 (5.0%) | 0 (0.0%) | 14 (2.3%) |

Data are number of participants and (percentages).

**Figure S1:**
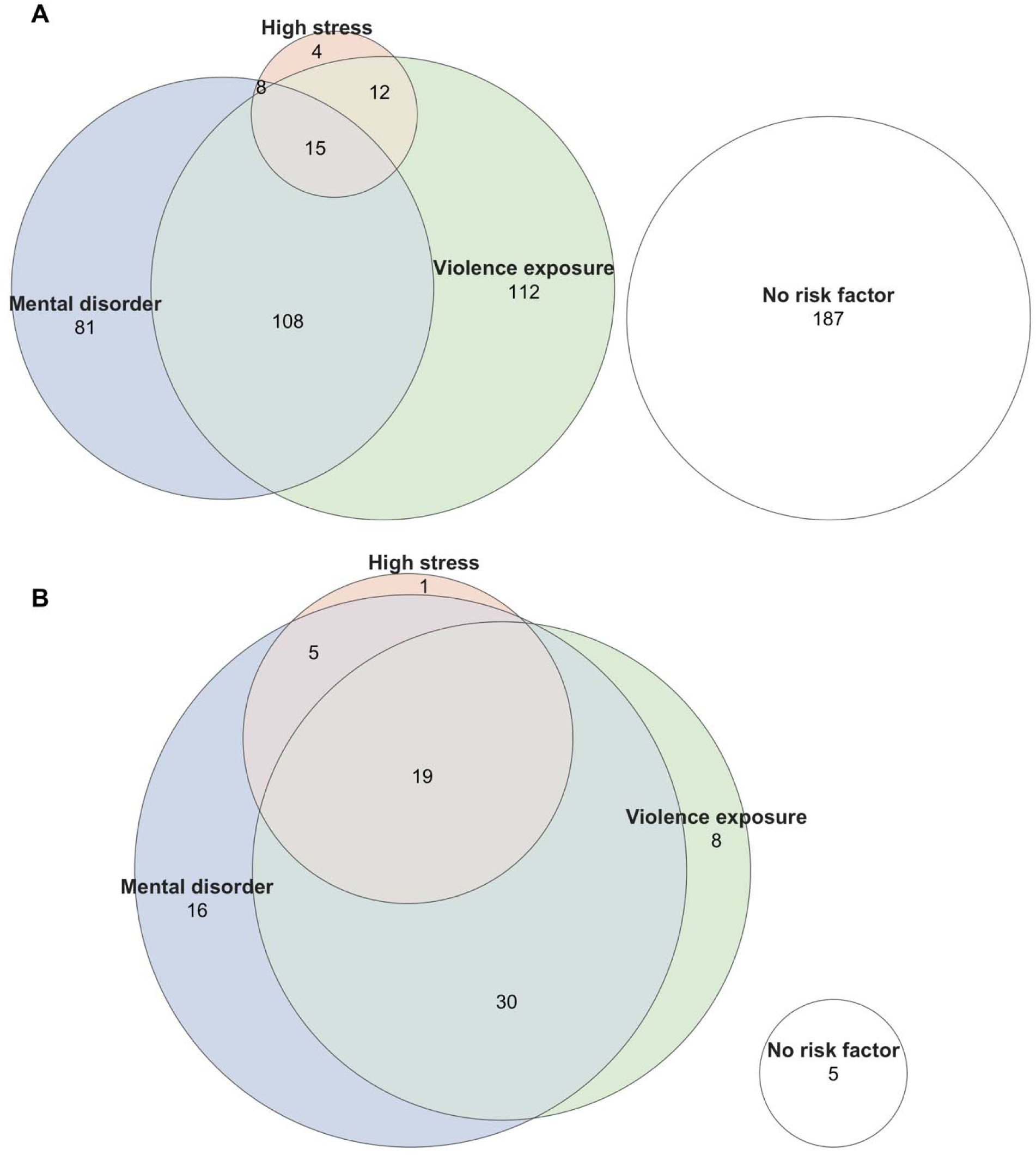
Co-occurrence of high perceived stress, violence exposure, and mental disorders. Panels: A) participants without suicidal ideation in the past 30 days, B) participants with suicidal ideation in the past 30 days. Areas represent approximate overlaps derived from Euler fits. Numbers indicate the number of participants in each intersection. In total, 527 participants without ideation and 86 participants with ideation were included. Mental disorder denotes the presence of any assessed current mental disorder. Violence exposure refers to lifetime exposure to violence in the family or community. High stress is defined as a Perceived Stress Score (PSS-4) ≥11. No risk factor indicates absence of all three risk factors. The intersection hig stress ∩ violence exposure without mental disorder (n = 2) among participants with ideation could not be represented by the Euler fit and is therefore not shown in panel B.

**Figure S2:**
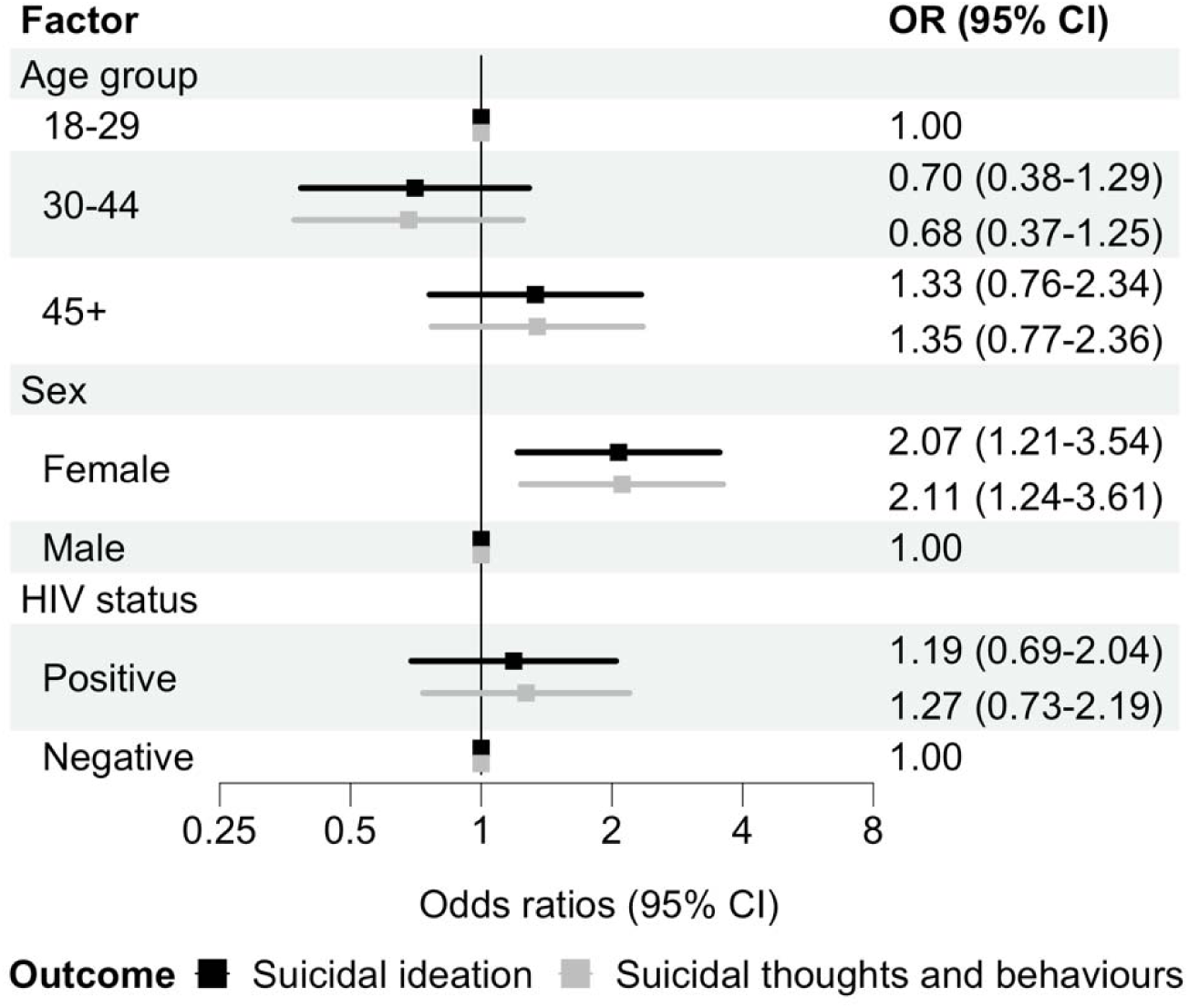
Associations between HIV status and past 30-day suicidal thoughts and behaviours. Adjusted odds ratios (ORs) for associations between HIV status and past 30-day suicidal ideation (black) and past 30-days suicidal thoughts and behaviours (grey). Models adjust for age group, sex, and population group. Error bars indicate 95% confidence intervals (CIs). Suicidal ideation is a binary outcome modelled with logistic regression. Suicidal thoughts and behaviours are coded hierarchically (none, ideation, plan, attempt), with participants assigned to the highest level endorsed, and modelled with ordinal logistic regression.

**Figure S3:**
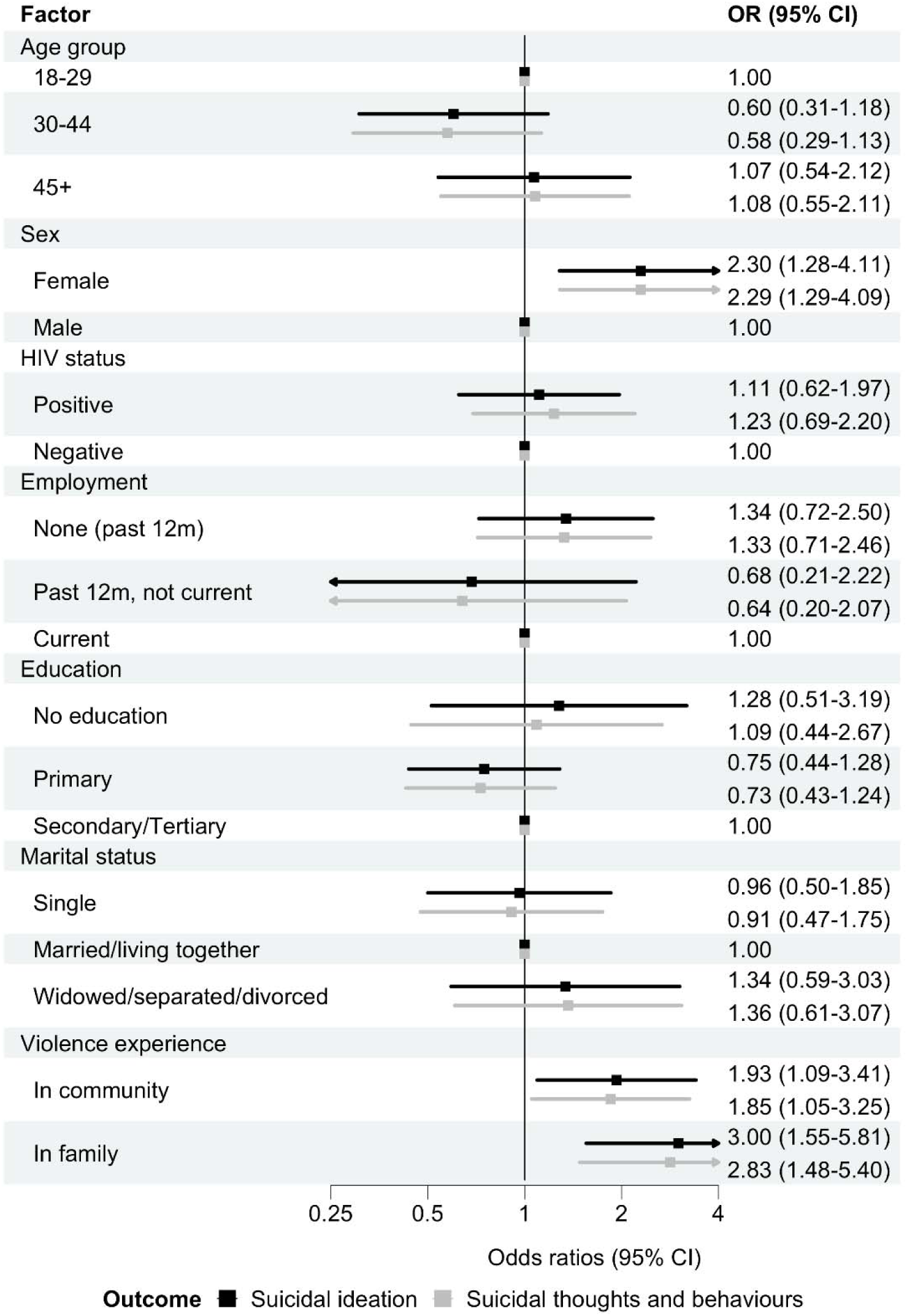
Sociodemographic factors and violence exposure associated with past 30-day suicidal thoughts and behaviours. Adjusted odds ratios (ORs) for associations between sociodemographic factors and violence exposure and past 30-day suicidal ideation (black) and past 30-days suicidal thoughts and behaviours (grey). Models included all sociodemographic factors shown, population group, and HIV status. Error bars indicate 95% confidence intervals (CIs). Suicidal ideation is a binary outcome modelled with logistic regression. Suicidal thoughts and behaviours are coded hierarchically (none, ideation, plan, attempt), with participants assigned to the highest level endorsed, and modelled with ordinal logistic regression.

**Figure S4:**
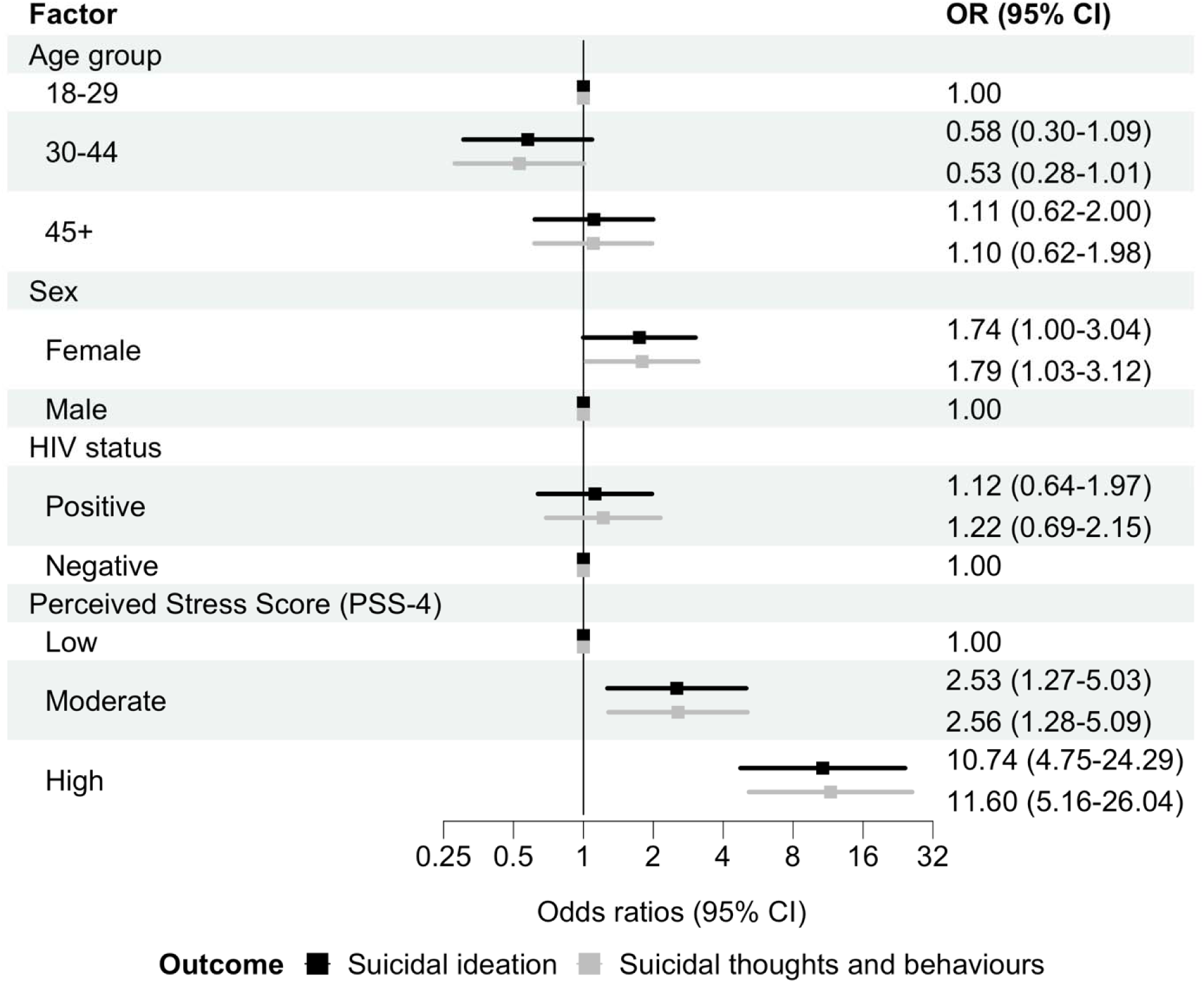
Associations between perceived stress and past 30-day suicidal thoughts and behaviours. Adjusted odds ratios (ORs) for associations between perceived stress and past 30-day suicidal ideation (black) and past 30-days suicidal thoughts and behaviours (grey). Models adjusted for age group, sex, population group, and HIV status. Error bars indicat 95% confidence intervals (CIs). Suicidal ideation is a binary outcome modelled with logistic regression. Suicidal thoughts and behaviours are coded hierarchically (none, ideation, plan, attempt), with participants assigned to the highest level endorsed, and modelled with ordinal logistic regression.

**Figure S5:**
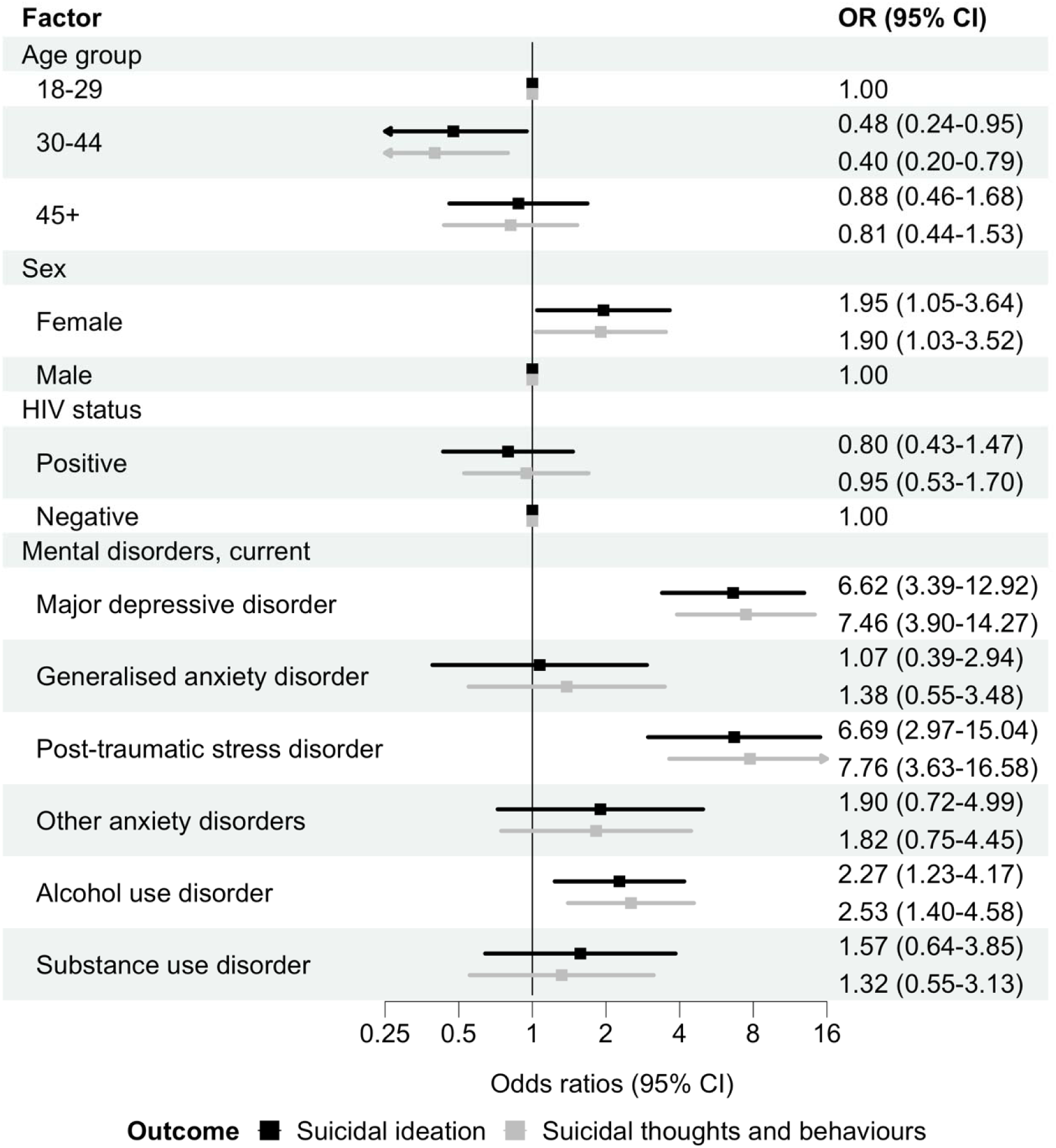
Associations between specific mental disorders and past 30-day suicidal thoughts and behaviours. Adjusted odds ratios (ORs) for associations between any current mental disorder and past 30-day suicidal ideation (black) and past 30-days suicidal thoughts and behaviours (grey). Models adjusted for age group, sex, population group, and HIV status. Error bars indicate 95% confidence intervals (CIs). Suicidal ideation is a binary outcome modelled with logistic regression. Suicidal thoughts and behaviours are coded hierarchically (none, ideation, plan, attempt), with participants assigned to the highest level endorsed, and modelled with ordinal logistic regression.

**Figure S6.**
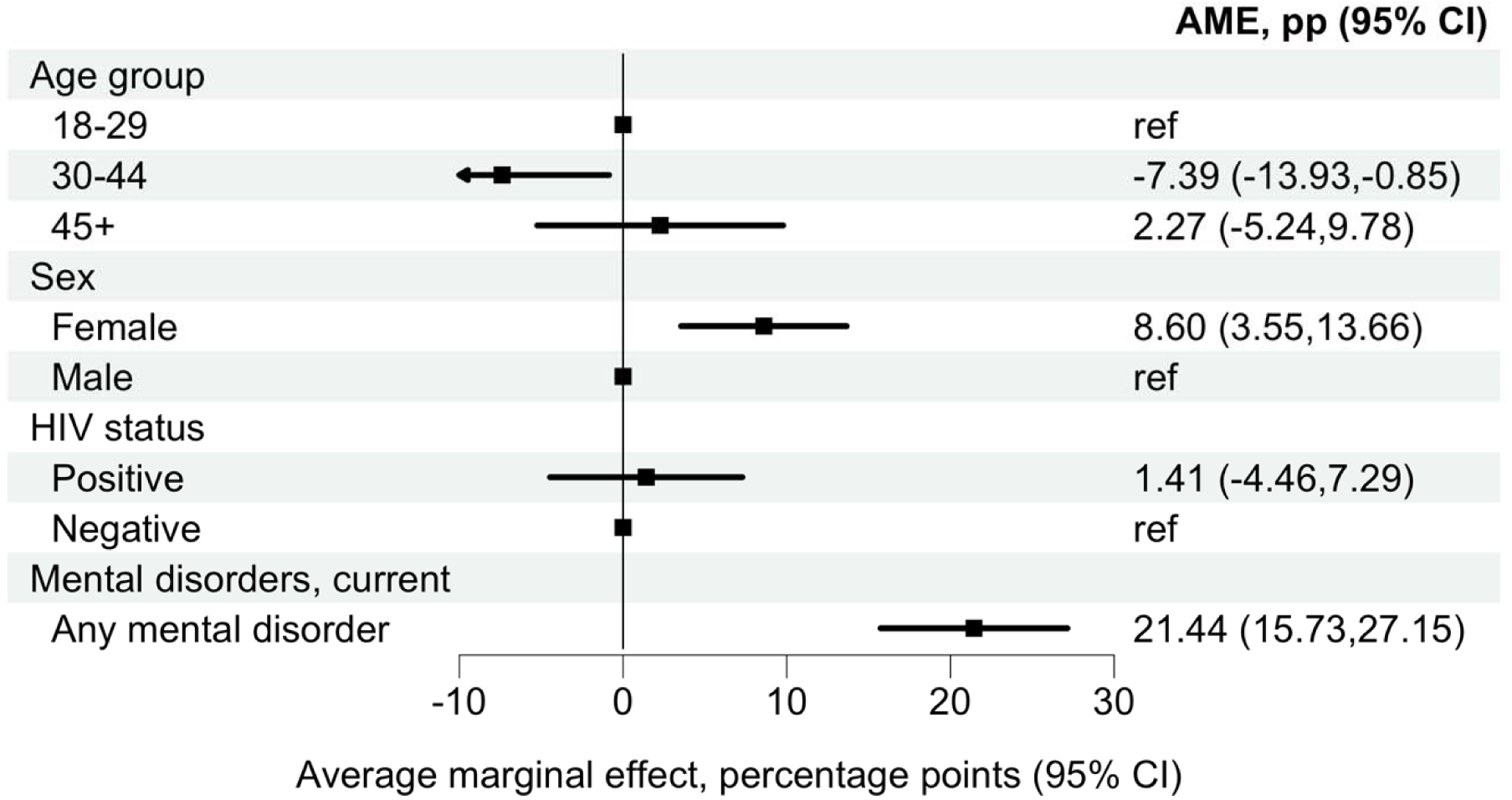
Average marginal effect of any current mental disorder on past 30-day suicidal ideation. Average marginal effect (AME), expressed as a percentage-point difference, for the association between any current mental disorder and past 30-day suicidal ideation. Estimates derive from an adjusted logistic regression model including age group, sex, population group, and HIV status. The AME represents the average discrete change in the predicted probability of suicidal ideation comparing participants with any current mental disorder versus none, averaged over the observed covariate distribution. Error bars indicate 95% confidence intervals. The corresponding adjusted odds ratio are presented in Figure S7.

**Figure S7:**
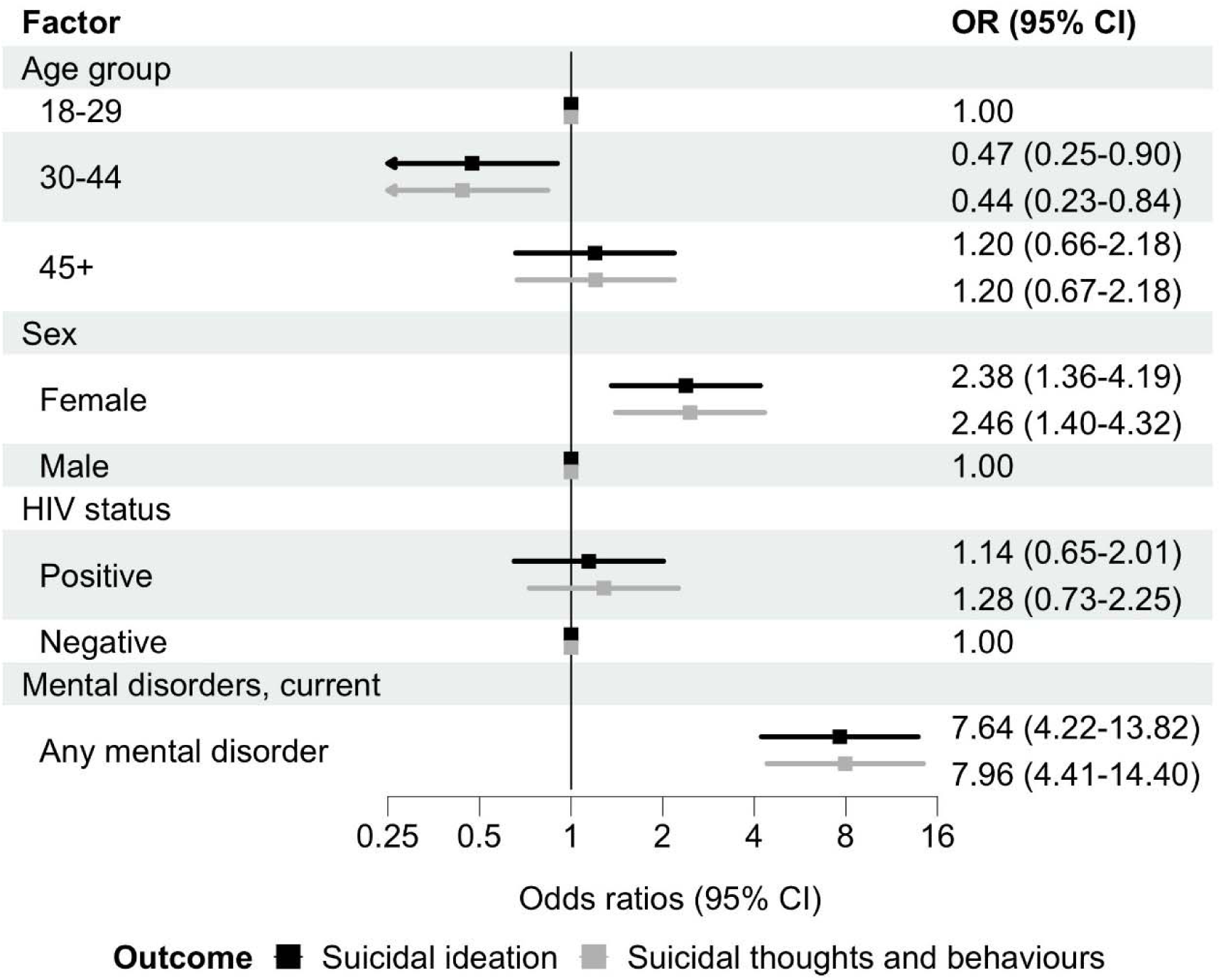
Associations between any mental disorder and past 30-day suicidal thoughts and behaviours. Adjusted odds ratios (ORs) for associations between any current mental disorder and past 30-day suicidal ideation (black) and past 30-days suicidal thoughts and behaviours (grey). Models adjusted for age group, sex, population group, and HIV status. Error bars indicate 95% confidence intervals (CIs). Suicidal ideation is a binary outcome modelled with logistic regression. Suicidal thoughts and behaviours are coded hierarchically (none, ideation, plan, attempt), with participants assigned to the highest level endorsed, and modelled with ordinal logistic regression.

**Figure S8.**
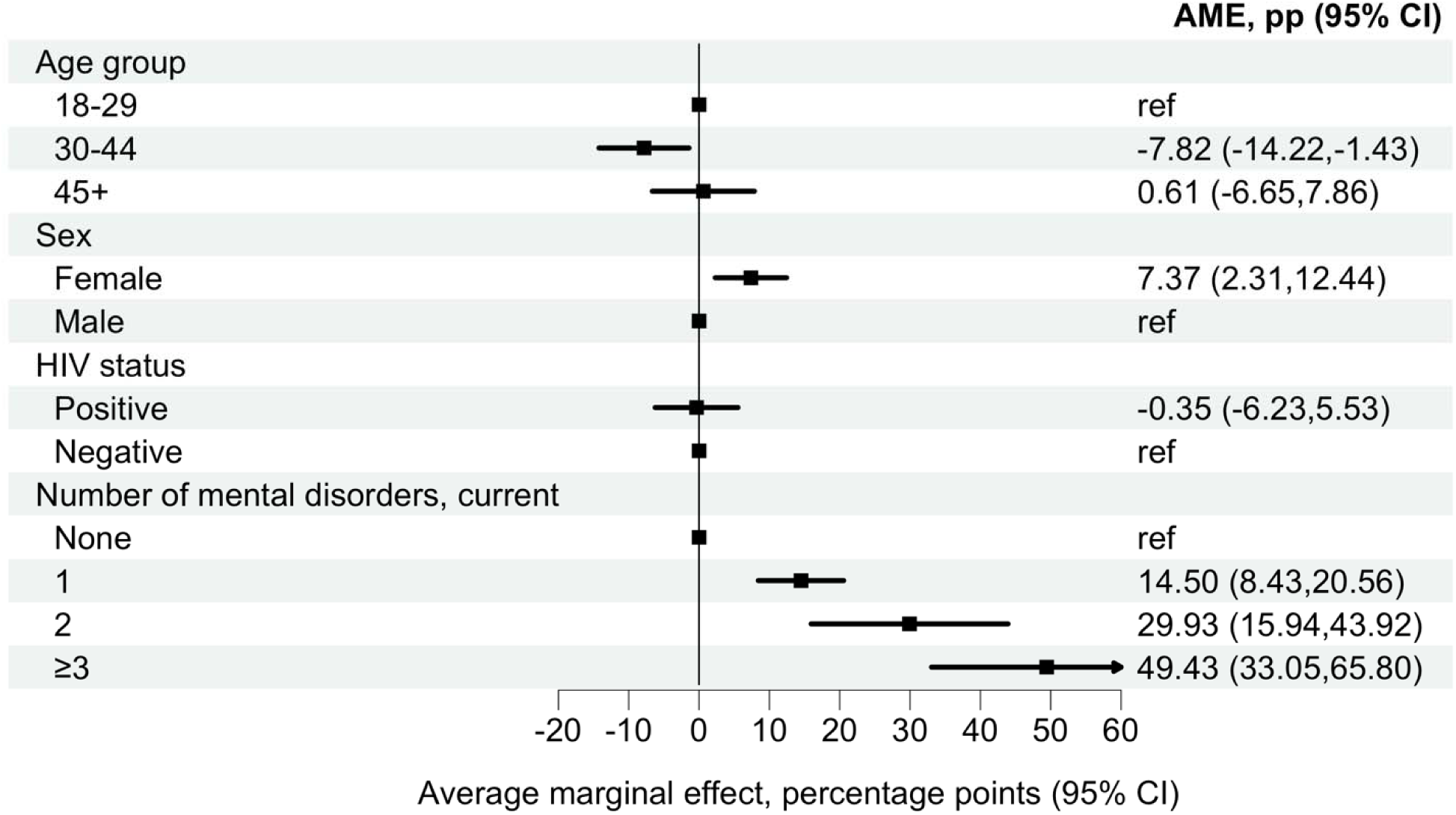
Average marginal effect of the number of current mental disorder on past 30-day suicidal ideation. Average marginal effects (AMEs), expressed as percentage-point differences, for the association between the number of current mental disorders and past 30-day suicidal ideation. Estimates derive from an adjusted logistic regression model including age group, sex, population group, and HIV status. AMEs represent the average discrete change in the predicted probability of suicidal ideation for each category of number of current mental disorders (1, 2, or ≥3) compared with none, averaged over the observed covariat distribution. Error bars indicate 95% confidence intervals. The corresponding adjusted odds ratio are presented in Figure S9.

**Figure S9:**
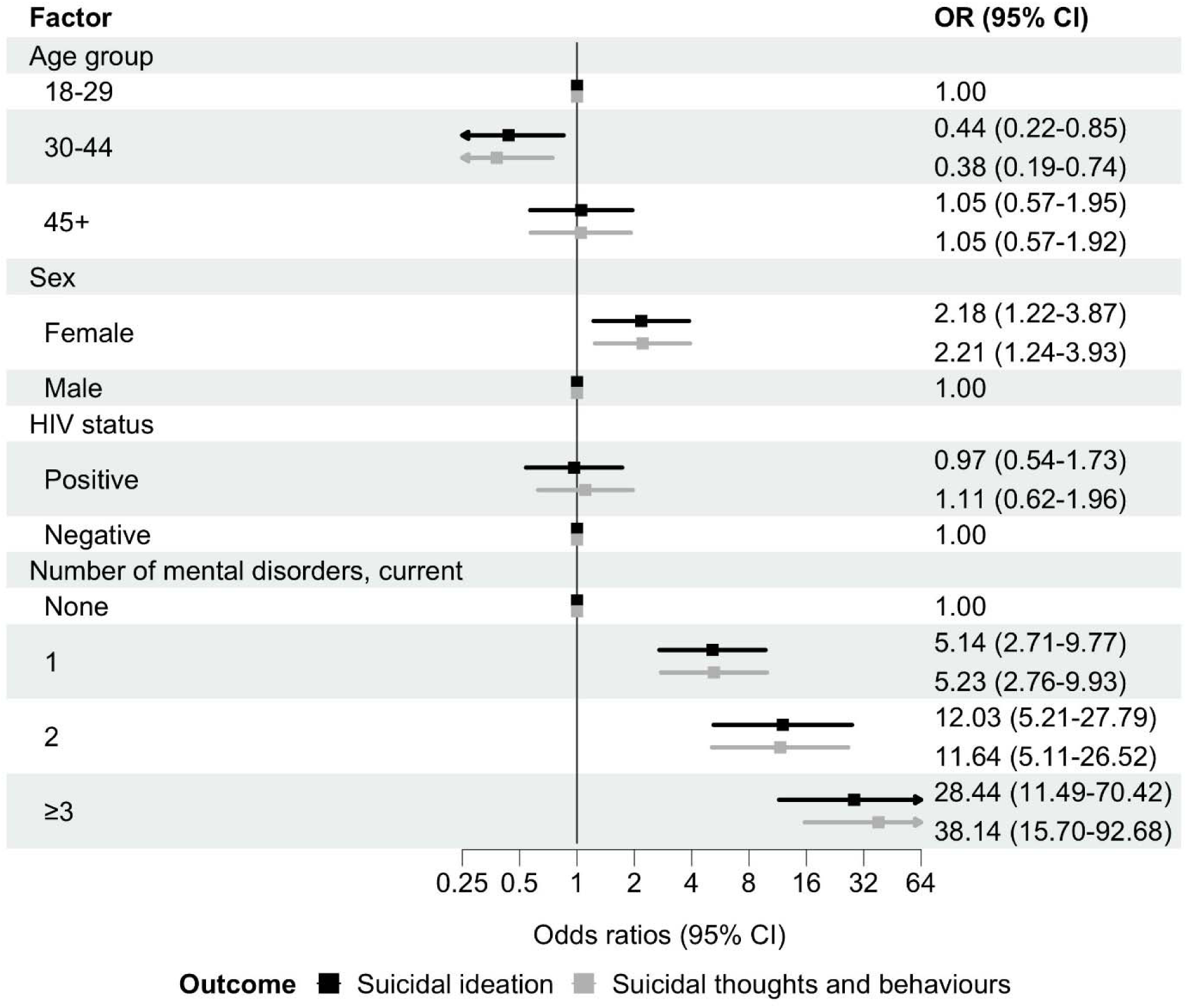
Associations between number of mental disorders and past 30-day suicidal thoughts and behaviours. Adjusted odds ratios (ORs) for associations between number of mental disorders and past 30-day suicidal ideation (black) and past 30-days suicidal thoughts and behaviours (grey). Models adjusted for age group, sex, population group, and HIV status. Error bars indicate 95% confidence intervals (CIs). Suicidal ideation is a binary outcome modelled with logistic regression. Suicidal thoughts and behaviours are coded hierarchically (none, ideation, plan, attempt), with participants assigned to the highest level endorsed, and modelled with ordinal logistic regression.

**Figure S10:**
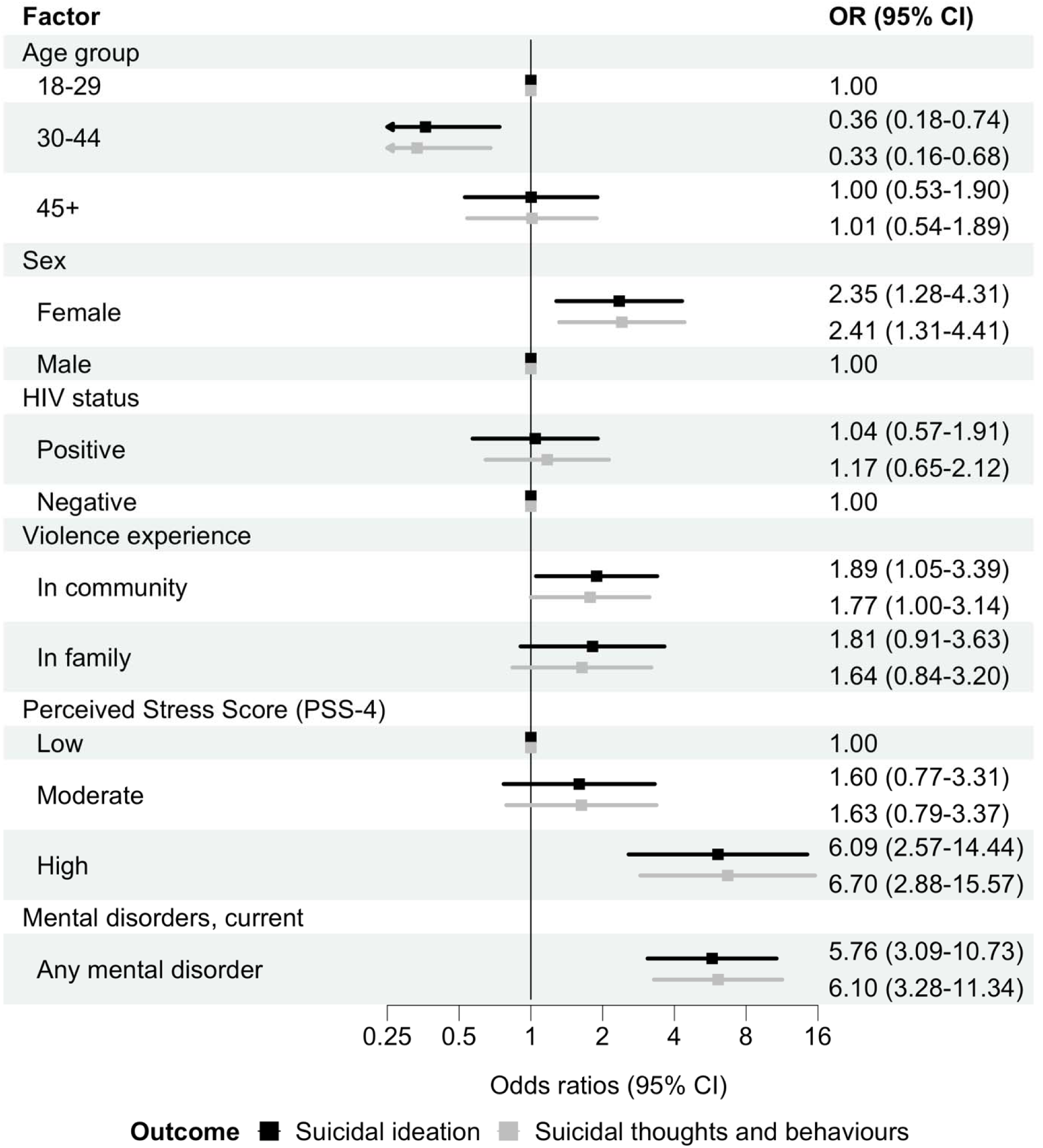
Associations between perceived stress and violence exposure and past 30-day suicidal thoughts and behaviours, adjusted for mental disorders. Adjusted odds ratios (ORs) for associations between perceived stress, and violence experience and past 30-day suicidal ideation (black) and past 30-days suicidal thoughts and behaviours (grey). Models adjusted for age group, sex, population group, HIV status, and the presence of any current mental disorder. Error bars indicate 95% confidence intervals (CIs). Suicidal ideation is a binary outcome modelled with logistic regression. Suicidal thoughts and behaviours are coded hierarchically (none, ideation, plan, attempt), with participants assigned to the highest level endorsed, and modelled with ordinal logistic regression.

**Figure S11:**
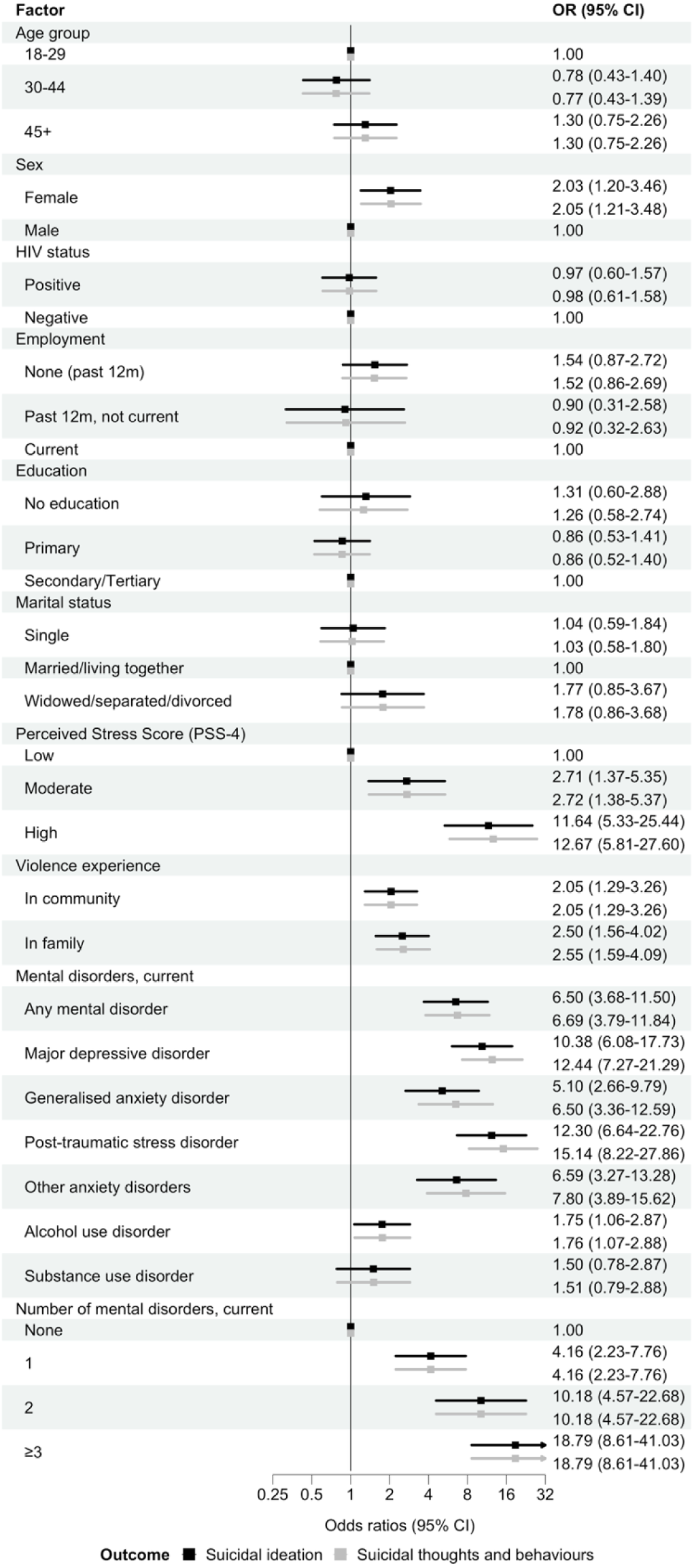
Unadjusted odds ratios for factors associated with past 30-day suicidal thoughts and behaviours. Unadjusted odds ratios (ORs) for factors associated with past 30-day suicidal ideation (black) and past 30-days suicidal thoughts and behaviours (grey). Error bars indicate 95% confidence intervals (CIs). Suicidal ideation is a binary outcome modelled with logistic regression. Suicidal thoughts and behaviours are coded hierarchically (none, ideation, plan, attempt), with participants assigned to the highest level endorsed, and modelled with ordinal logistic regression.

